# Phase-wise Impact Analysis of the Indian National Lockdown against COVID-19 Outcomes

**DOI:** 10.1101/2022.10.27.22281585

**Authors:** Vishwali Mhasawade, Siddhesh Zadey, Aatmika Nair

## Abstract

India was one of the most vulnerable countries to the COVID-19 pandemic considering the high transmissibility of the virus, exploding population, and fragile healthcare infrastructure. As an early counter, India implemented a country-wide lockdown and we aimed to study the impact of 4 lockdowns and 2 unlock phases on 6 outcomes: case growth, death count, effective reproduction number, mobility, hospitalization, and infection growth by two methods: interrupted time series (ITR) analysis and Bayesian causal impact analysis (BCIA) for nationals and sub-national levels. We observed that the effects are heterogeneous across outcomes and phases. For example, ITR revealed the effect to be significant for all the outcomes across all phases except for case growth in phase 1. BCIA revealed that the causal effect of all four lockdown phases was positive for deaths. At the state level, Maharashtra benefited from the lockdown in comparison to Tripura. Effects of lockdown phases 3 and 4 on death count were correlated (R=0.70, p<0.05) depicting the ‘extended impact’ of phase-wise interventions. We observed the highest impact on mobility followed by hospitalization, infection growth, effective reproduction number, case growth, and death count. For optimal impact, lockdown needs to be implemented at the sub-national level considering various demographic variations between states.

## Main

Severe Acute Respiratory Syndrome Coronavirus-2 (SARS-Cov-2) is a novel virus causing Coronavirus Disease 2019 (COVID-19). The first observed cases of ‘pneumonia of unknown cause’ emerged in Wuhan, China in December 2019. Since then it has infected 548,868,275 people and caused 6,338,655 deaths globally till June 2022^1^. SARS-Cov-2 transmits through multiple modes such as human-to-human contact and droplets^2^, necessitating non-pharmaceutical interventions (NPI) such as physical distancing, quarantine, and isolation, among others in the early phase of the disease spread in the absence of vaccines and treatments^3^.

As early mitigation strategies in response to the outbreak, social distancing measures and travel restrictions were adopted in China^4^. ‘Unprecedented in public health history’, the Wuhan and Hubei regions were put under ‘lockdown’ on 23rd and 24th January 2020, respectively, effectively^5^. This lockdown included a travel ban in and out of these cities, schools, and entertainment areas were closed and public gatherings were prohibited. Haider et al have arbitrarily defined lockdown as ‘a set of measures aimed at reducing transmission of COVID-19 that are mandatory, applied indiscriminately to a general population and involve some restrictions on the established pattern of social and economic life^6^. Due to COVID-19’s pandemic status, in March 2020, around 149 countries including India adopted physical distancing, school, and workplace closures, restrictions on mass gatherings, public transport, and lockdowns^7^. By mid-April 2020, over 3.9 billion people globally, were under complete or partial lockdown^8^. The purpose of stringent movement restrictive public health NPI such as lockdown is to slow the viral transmission for the health system to scale up and enhance its preparedness^9^. Early on, some natural experiments and observational studies found positive evidence for lockdown as an intervention against COVID-19 case incidence^10^ and mortality^11^.

In India, the first COVID-19 case was reported on 30^th^ January 2020 in Thrissur, Kerala, for a 20-year-old female who had returned from Wuhan, China^12^. Considering the high transmissibility^13^, a large and densely-packed population of over 1.3 billion, diverse cultural values, socio-economic disparities, health equalities, and a fragile healthcare infrastructure^14,15^, India was perceived to be one of the most vulnerable regions in the world^16^. It was estimated that without any public health interventions, India would have 2.2 million cases as of 15^th^ May 2020 as opposed to 13,800 observed cases under a stringent public health intervention^17^. Hence, early lockdown implementation was considered necessary to curb the devastation due to the pandemic.

In early March 2020, India adopted containment strategies including quarantining individuals from high case burden countries, isolating infected individuals, restricting the local movement of people in high-incidence areas^18^, closing schools and workplaces, canceling mass gatherings, etc.^19^. On 22^nd^ March 2020, a ‘*Janta Curfew*’ (voluntary public curfew) was observed followed by one of the largest and most extended country-wide lockdowns expanding over 4 phases starting from 25^th^ March 2020. India’s swift implementation of lockdown was praised by WHO as ‘tough and timely’^15^. The lockdown phases differed by stringency^20^, functioning of businesses, travel restrictions, and zone classification of districts based on case burdens^17^. The first lockdown phase was from 25^th^ March to 14^th^ April 2020 (21 days), the second phase was from 15^th^ April to 3^rd^ May 2020 (19 days), the third phase was from 4^th^ to 17^th^ May 2020 (14 days), and the fourth lockdown phase was from 18^th^ to 31^st^ May 2020 (14 days). Post-lockdown re-opening (or unlocking) was also gradual with the first unlock phase being from 1^st^ to 30^th^ June 2020 (30 days), while the second unlock phase was from 1^st^ to 31^st^ July 2020 (31 days)^21^. The first unlock phase saw lockdown only in containment zones while shops, hotels, and restaurants were started in a phased manner in other zones. The second unlock phase permitted most activities to function, inter and intrastate travel was opened as well as limited international travel was permitted under Vande Bharat Mission^22^.

Previously, several epidemiological modeling studies have predicted the number of COVID-19 cases and deaths under diverse scenarios hypothesizing the presence and space of lockdown. However, only a few studies have investigated the effectiveness of lockdown phases based on observed data^10^. A national-level impact analysis would be insufficient to identify how the phases affected COVID-19-related outcomes. Hence, it is necessary to assess the efficacy of the lockdown at a lower, i.e. subnational geographic and administrative levels. We aimed to study the impact of the four lockdown phases, and the 2 unlock phases on four COVID-19 outcomes 1) death rate, 2) case growth, 3) effective (time-varying) reproduction number (R_t_), and 4) mobility at the national (all India) and sub-national level using two analytical methods: interrupted time-series regression (ITR) and Bayesian causal impact analysis. Our findings provide evidence for the effectiveness of early public health interventions such as lockdowns to control the pandemic and facilitate a deeper and policy-oriented understanding of the COVID-19 threat in a densely populated country like India and its states.

## Results

With the ITR analysis, we observed heterogeneity in the models across outcomes and phases. The supplementary material **(Table S1)** presented the best-performing model considering the multi-level nature of the data using ITR analysis for the different outcomes and lockdown and unlock phases which are followed by their 95% confidence intervals and significance values. Interesting to note that the lockdown intervention did not have a significant effect on case growth in phase 1 but was significant across later phases as outlined in **Table 2**. However, we observed that for the rest of the lockdown and unlock phases the interventions did have a significant impact on all the outcomes, but the magnitude of the effect is heterogeneous with the highest effect on mobility and hospitalization. Furthermore, we observed that the magnitude of the effect of lockdown interventions on mobility and hospitalization outcomes was around 7 times that of other outcomes. Considering the multi-level nature of the data, details about the random and fixed effects of the different outcomes and phases can be found in the supplementary material **(Tables S4-S39)**. Furthermore, we observed that the random effects are significant with considerable variance at the subnational levels as demonstrated in Tables S4-S39. This suggests that we need to analyze the effects at the subnational level rather than considering an aggregate model.

**Table 1:** Model performance (AIC) of ITR for different outcomes across phases.

| Phase | Case growth | Death count | Rt | Mobility | Hospitalization | Infection Growth |
| --- | --- | --- | --- | --- | --- | --- |
| Lockdown phase 1 | -5068.784 | 36079.118 | 25177.188 | 33713.709 | 69213.208 | -7353.731 |
| Lockdown phase 2 | -5017.813 | 36064.055 | 25168.100 | 33665.835 | 69208.942 | -7380.255 |
| Lockdown phase 3 | -5032.557 | 36064.979 | 25185.604 | 34154.529 | 69214.503 | -7369.133 |
| Lockdown phase 4 | -5025.311 | 36082.749 | 25195.585 | 33964.615 | 69211.559 | -7346.045 |
| Unlock phase 1 | -5016.209 | 36027.696 | 25188.474 | 32850.881 | 69195.283 | -7347.558 |
| Unlock phase 2 | -5017.825 | 36022.442 | 25178.963 | 34122.322 | 69138.579 | -7357.981 |

**Table 2:** **Impact of interventions on COVID-19 outcomes as assessed using Interrupted time series analysis, effect [95% CI] (significance), note that significance symbols are only represented when the p-value is significant. Rows represent the different phases of lockdown and unlock; columns represent the different outcomes. The model equations for each phase and outcome are mentioned in the supplementary Tables S4-S39.**

| Phase | Case growth | Death count | Rt | Mobility | Hospitalization | Infection Growth |
| --- | --- | --- | --- | --- | --- | --- |
| Lockdown phase 1 | -0.001<br>[-0.002<br>,0.001] | 0.385<br>[0.329<br>,0.440]<br>(***) | 0.046<br>[0.012<br>,0.079]<br>(**) | 7.176<br>[7.023<br>,7.328]<br>(***) | 8.143<br>[6.249<br>,10.036] (***) | 0.003<br>[0.001<br>,0.004] (***) |
| Lockdown phase 2 | 0.001<br>[-3.01E-05<br>,0.002] (.) | 0.308<br>[0.257<br>,0.360]<br>(***) | 0.0840<br>[0.052<br>,0.115]<br>(***) | 7.365<br>[7.220<br>,7.512]<br>(***) | 6.457<br>[4.677<br>,8.237] (***) | 0.004<br>[0.003<br>,0.005] (***) |
| Lockdown phase 3 | 0.002<br>[0.001,<br>0.003] (**) | 0.321<br>[0.271<br>,0.371]<br>(***) | 0.063<br>[0.032<br>,0.094]<br>(***) | 7.775<br>[7.624<br>,7.926]<br>(***) | 6.985<br>[5.252<br>,8.718] (***) | 0.004<br>[0.003<br>,0.005] (***) |
| Lockdown phase 4 | 0.002<br>[0.001<br>,0.003] (**) | 0.339<br>[0.290<br>,0.389]<br>(***) | 0.060<br>[0.030<br>,0.912]<br>(***) | 7.854<br>[7.706<br>,8.002]<br>(***) | 7.019<br>[5.300<br>,8.738] (***) | 0.003<br>[0.002<br>,0.005] (***) |
| Unlock phase 1 | 0.001<br>[0.001<br>,0.003] (*) | 0.339<br>[0.290<br>,0.388]<br>(***) | 0.0655<br>[0.036<br>,0.097]<br>(***) | 7.899<br>[7.677<br>,8.030]<br>(***) | 7.318<br>[5.610<br>,9.026] (***) | 0.0036<br>[0.0024<br>,0.0004] (***) |
| Unlock phase 2 | 0.001<br>[4.08E-05<br>,0.004] (*) | 0.2630<br>[0.211<br>,0.315]<br>(***) | 0.067<br>[0.036<br>,0.098]<br>(***) | 7.792<br>[7.642<br>,7.942]<br>(***) | 4.564<br>[2.763<br>,6.366] (***) | 0.004<br>[0.003<br>,0.005] (***) |

Using Bayesian causal impact analysis, we evaluated the effect of the interventions through the absolute causal effect values obtained using the causal impact analysis, while also performing a sensitivity analysis of -3 to +3 days around the intervention times. The analysis at the national level is reported in **Table 3**, with each cell denoting the causal effect of the intervention at the national level. We observed that for case growth the interventions during lockdown phases 1, 2, 3, and 4 had an overall negative effect, indicating unlock phases 1 and 2 had a positive causal effect on case growth. We also found that lockdown phases 1, 2, 3, and 4 had an overall positive causal effect on death rate and mobility while unlock phase 1 had a positive effect on death rate and mobility, while unlock phase 2 had a positive effect on death rate but not on mobility. Lockdown phases 1, 2, and 3 had a positive effect on R_t_ but phase 4 had a negative effect. Unlock phase 1 had a positive effect on R_t_ while phase 2 had a negative effect. Thus, even at the national level, we observed a lot of heterogeneity in the effects across phases and outcomes. These (normalized) differences are even more apparent at a subnational level reflected in **Figure 1**. For example, for the union territory of Jammu and Kashmir, there was an increase in the effect on case growth across lockdown phases 1, 2, 3, and 4 while the effect decreased across unlock phases 1 and 2. However, in the same union territory, there was an overall decrease in the effect on death rate across lockdown phases 1, 2, 3, and 4 while the effect remained almost constant during unlock phases 1 and 2. In the state of Gujarat, the effects on R_t_ gradually increased across lockdown phases 1, 2, 3, and 4. In the state of Jharkhand, there was a higher effect on mobility during lockdown phase 1 which was considerably higher than other neighboring states and union territories; this effect remains almost constant across lockdown phases 2, 3, and 4. In general, the interventions had a heterogeneous effect on states in any intervention period and this also changes across outcomes.

**Table 3:** **Absolute causal effect of intervention (with 95% confidence interval) as computed using causal impact analysis for different outcomes. Rows represent different intervention phases and columns represent the outcomes.**

|  | Outcome (95% Credible Interval) |  |  |  |  |  |
| --- | --- | --- | --- | --- | --- | --- |
| Phase | Case growth | Death count | $R_t$ | Mobility | Infection Growth | Hospitalization |
| Lockdown phase 1 | -0.465<br>[-1.064, 0.059] | 0.001<br>[0.000, 0.001] | -0.139<br>[-0.224, -0.055] | 9.223<br>[-13.262, 29.495] | 0.042<br>[0.007, 0.075] | 180.53 [-19.300, 376.350] |
| Lockdown phase 2 | -0.078<br>[-0.362, 0.211] | 0.002<br>[0.002, 0.003] | 0.363<br>[0.268, 0.461] | 38.156<br>[24.474, 51.235] | 0.044<br>[0.023, 0.064] | 17.331 [-209.130, 239.710] |
| Lockdown phase 3 | -0.014<br>[-0.247, 0.189] | 0.002<br>[0.001, 0.003] | 0.209<br>[0.115, 0.301] | 28.231<br>[17.679, 38.909] | 0.0198<br>[0.001, 0.036] | 587.09 [335.800, 834.790] |
| Lockdown phase 4 | -0.002<br>[-0.222, 0.177] | 0.007<br>[0.006, 0.009] | 0.195<br>[0.109, 0.278] | 24.975<br>[15.161, 35.250] | 0.013 [-0.001, 0.029] | 1157.90 [838.180, 1479.220] |
| Unlock phase 1 | 0.004<br>[-0.153, 0.170] | 0.006<br>[0.003, 0.008] | 0.175<br>[0.044, 0.299] | 24.571<br>[15.630, 33.406] | 0.030<br>[0.017, 0.044] | 2641.270 [2216.410, 3056.71] |
| Unlock phase 2 | 0.008<br>[-0.115, 0.122] | 0.012<br>[0.010, 0.015] | 0.066<br>[-0.049, 0.203] | -1.057<br>[-8.780, 6.459] | -0.004 [-0.015, 0.005] | 5613.82 [4333.120, 6857.320] |

**Figure 1:**
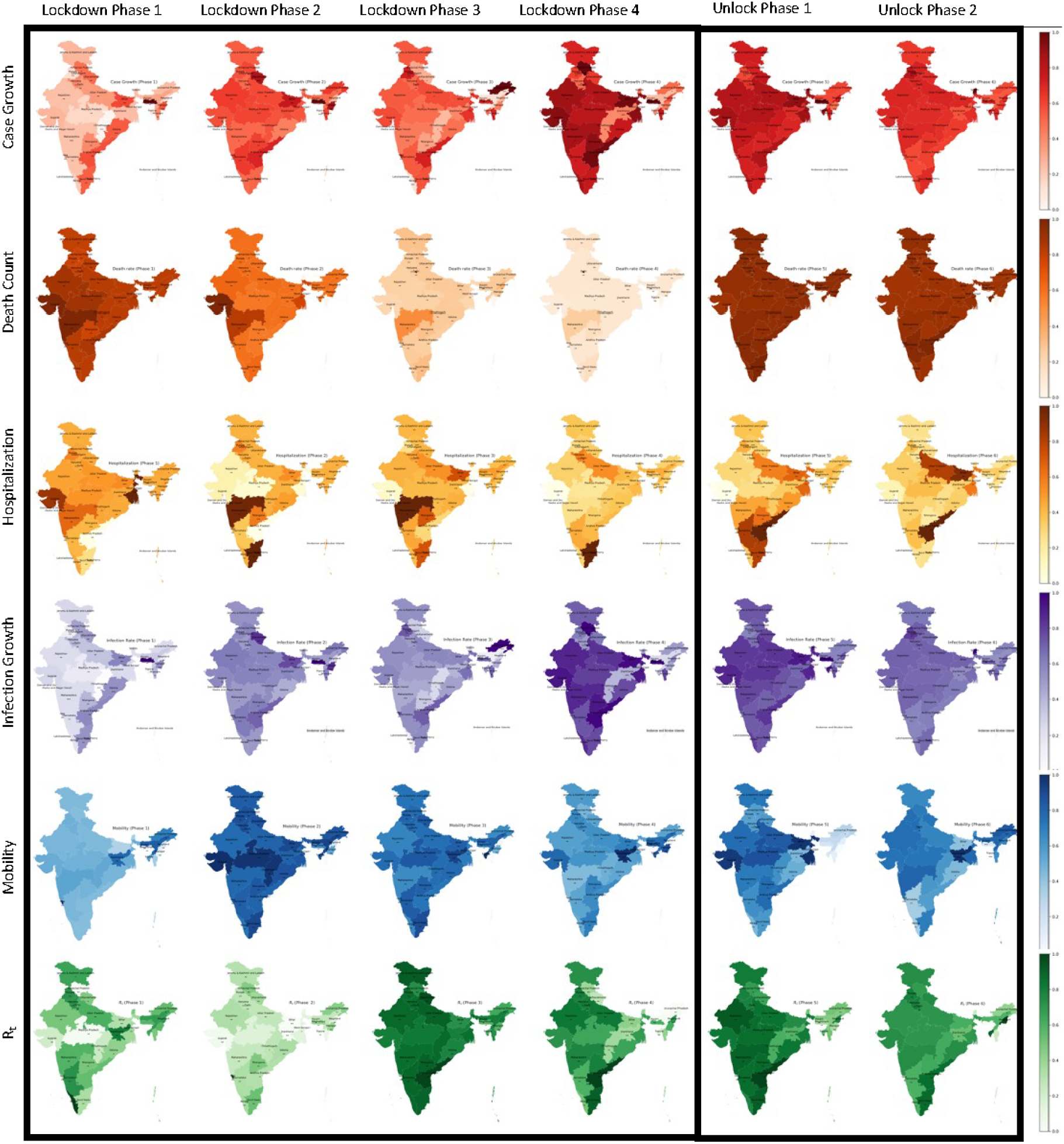
**Effects of lockdown and unlock interventions as estimated using causal impact analysis for different outcomes (rows) across phases (columns) for Indian states and UTs. For lockdown phases 1 to 4 (first 4 columns) values closer to 1 represent a higher effect of the intervention which is desired, while for the unlock interventions (columns 5 and 6) values closer to 0 are desired. Darker colors denote values closer to 1 while lighter colors denote values closer to 0. States for which the causal effect for each different outcome was significant (p<0.05) are mentioned for each phase.**

To understand the lingering effects of the interventions, we also evaluated the spearman correlation between causal effects obtained using causal impact analysis across outcomes and lockdown and unlock phases. The correlation (blue for positive and red for negative magnitudes) between the effects across phases and outcomes was significant only in certain cases as represented by the white asterisk in **Figure 2**. Another interesting phenomenon was that we did not observe significant self-correlation between most of the effects. We also saw a significant positive correlation between the effects of death count and hospitalization indicating the severity of the cases that were hospitalized. Furthermore, we also observed a correlation between Rt and infection growth, indicating the causal chain between the two. These factors were important in assessing how the effects interact across outcomes for different phases. Thus, assessing the correlation between the effects across phases and outcomes allowed the identification of any long-term effect on one intervention, helpful in assessing long-term effects to guide efficient policy decisions.

**Figure 2:**
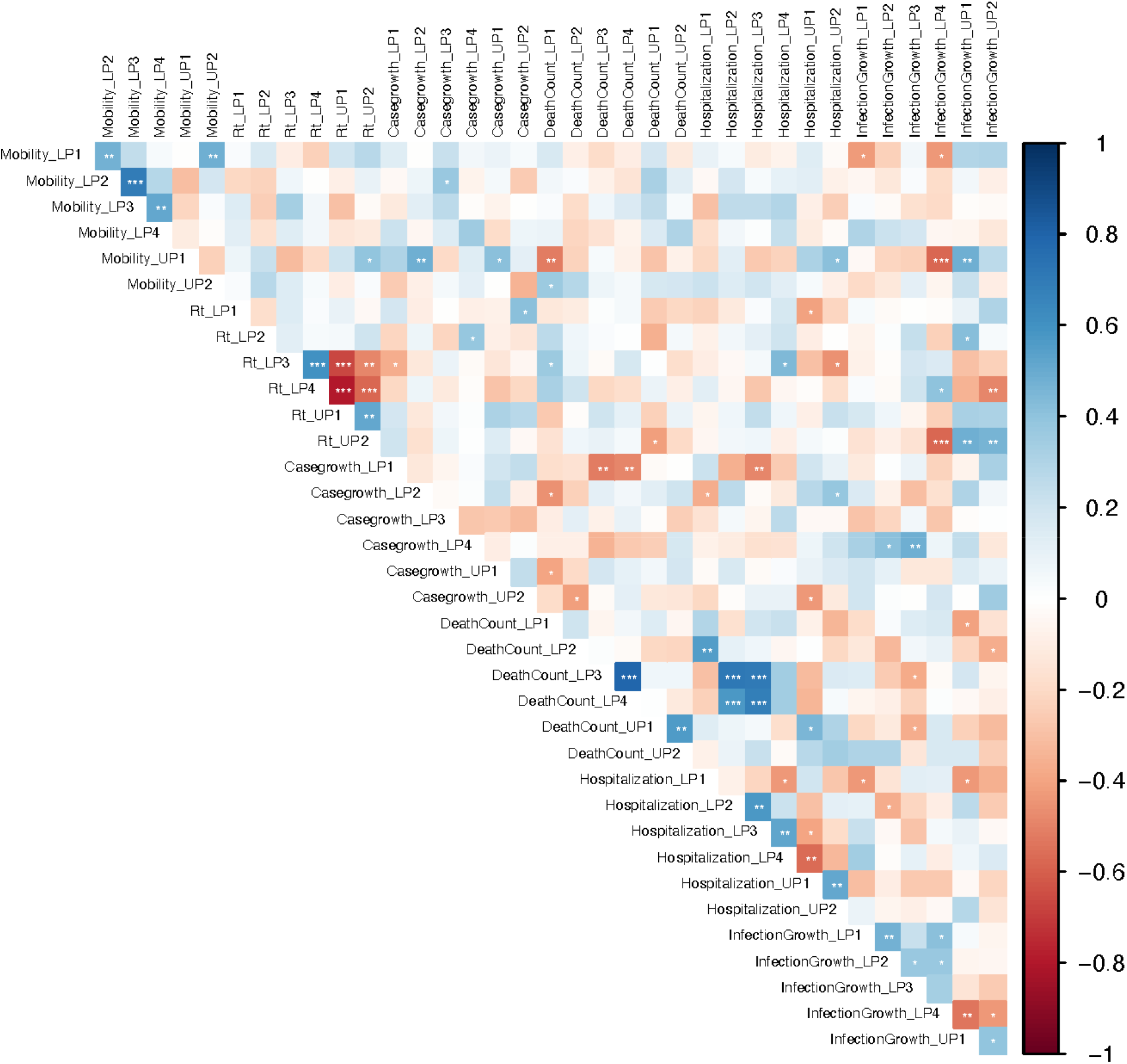
Correlation between causal impact effects across outcomes and phases.

## Discussion

Interrupted time series regression model results suggest that the effects varied significantly at the subnational level as evidenced by the multi-level modes, with the effect being significant on mobility and hospitalization across all phases. Interestingly, we observed that the lockdown interventions did not have a significant effect on case growth during lockdown phases 1 and 2 highlighting the effectiveness of such interventions in curbing case growth which was one of the fundamental outcomes to consider. Investigating the effect estimates for different phases and outcomes illustrates the variability at the sub-national or state level, motivating the causal impact analysis to be performed at this level. The Bayesian causal impact analysis illustrates how the effects of the interventions changed across phases for different outcomes. For example, for case growth, we observed that the effect for Jammu and Kashmir gradually increased across the lockdown phases but was almost constant in the unlock phases. Similarly, for the death count, we observed that the effect size for Maharashtra gradually decreases with the progression of the pandemic across the lockdown phases, also reflected in the hospitalizations. For infection growth, we saw almost the same effect across all phases in West Bengal while for Gujarat, a significant effect on mobility during the lockdown phases except phase 1. Kerala had a significant effect across all the phases. In the correlation analysis of outcomes across phases and places, we observe that the effects are correlated for certain phases and outcomes, but this is not significant in most of the combinations. For example, we found that effects on R_t_ are correlated across lockdown phases 3 and 4 and unlock phases 1 and 2. Moreover, we also found that the effect on the death count in lockdown phase 3 highly correlated with the effect on hospitalizations in lockdown phases 3 and 4. This was surprising considering that a higher effect on hospitalization should reduce the effect on death counts but this was not observed here.

Tiwari and colleagues through five compartment mathematical model Susceptible (S)-Exposed (E)-Infected (I)-Recovered (R)-Death (D) (SEIRD), investigated the progression of COVID-19 in India from 30th January to 10th July and the impact of lockdown on R_t_ and case growth^23^. Their findings show that the cases were fewer during the lockdown phases and have increased sharply during unlock phases. Our study concurs that the lockdown slowed down the case growth but unlocking spiked it. With respect to R_t_, Tiwari reported a general decrease in the trend during the lockdown period and an increase during the post-lockdown period. Whereas by the ITR method, we found that lockdown phases 1, 3, and 4 impeded R_t_, but lockdown phase 2 observed a spike. Moreover, unlock phases 1 & 2 showed a decrease slowed down the increase in R_t_. This shows that it is vital that we observe the effect of the lockdown in a phase-wise manner that was implemented in the country. Another study using the epidemic SIR model shows that within the lockdown phase 1, R_t_ showed a dip followed by a rise and then went down to 1.56^24^. Another study investigated R_t_ for 10 states in India at 15 days and 30 days after implementing the lockdown and showed that the highest decrease in R_t_ was seen in Andhra Pradesh, Delhi, and Rajasthan, and a reciprocal increase in R_t_ in Gujarat during the same period^25^. In our study too, R_t_ in Gujarat during lockdown phase 1 increased while it decreased in Kerala, Jharkhand, Punjab, and Delhi. A study done to investigate the impact of lockdown in the city of Pune, Maharashtra state showed that the regional lockdown showed a 13% decrease in weekly new patients while the national lockdown showed only a 2% decrease^26^. This validates the need to study the sub-national impact of lockdown on COVID-19 as done in our study.

Bihar, Karnataka, Andhra Pradesh, and Tamil Nadu observed an increase in death rate in the post-lockdown phases^27^, similar to our findings that showed an increase in death rates during the same period. By simple regression analysis, Goshal et al. found that there was a 45% reduction in total infection in India after one week from the declaration of the lockdown which conforms to our findings of case growth that slowly declined throughout the lockdown phases^28^. Thayer et al used Google community mobility reports which categorized mobility into six parts and showed that mobility at the parks, recreational areas, workplaces, and transit stations was reduced when the lockdown was announced and continued to remain low till unlock phase 1^29^. In our study, we found that Lockdown phases 1, 2, 3, and 4 had a negative effect on mobility whereas unlock phase 1 had a positive effect on mobility.

For most outcomes, the lockdown did not have a significant impact as illustrated using the causal impact analysis. One potential reason for this could be the lack of homogeneity at the national level with respect to healthcare resources and the impact on jobs. This suggests a better approach for addressing such settings while considering economic impact at a more regional level. A modeling study showed that a longer lockdown between 42-56 days is preferred to flatten the curve rather than 21 days lockdown^30^. However, lockdown phases 3 and 4 observed relaxations of some norms in certain zones, but our study did not observe much difference in these phases. Moreover, there is geographic variability in the effects of implementing the policy (non-monolithic structure of the states), and hence a bottom-up approach is desired in designing interventions in comparison to top-down approaches even in the initial phases of the pandemic. Thus, there is a need to equally involve state and local governments. We further observed that there was excess focus on the anticipated efficacy of the interventions rather than to scale up the healthcare resources, and thus preparedness for the impact of the pandemic was lost. This was more of a speculated efficacy, and thus, very short-sighted policy implications with little consideration of societal implications and ethical concerns of the lockdown interventions^31^.

There are certain limitations of this work. For example, some outcomes such as epidemiological doubling time are not included. For the current outcomes considered it is challenging to estimate the accuracy of the values, hence we use the processed data from IHME. To address this higher resolution data, maybe at the district level would be helpful but is currently not possible due to 1) very limited data for the specific time period^32^, and 2) very few districts were infected during the lockdown periods. Another major limitation is the simplicity of the Bayesian causal impact model with no time-varying factors, and no confounders that can potentially generate better counterfactuals. However, this is because 1) the predictor needs to be correlated to the outcomes, and 2) the predictor should not be impacted by the intervention. As we did not find such predictors that satisfy the second condition, we are restricted to a simple model. For future work, predictors from other places such as Sweden and Korea where interventions were not implemented during our study period could be incorporated.

## Methods

### Data Sources

We used COVID-19 daily projections for six outcomes - 1) mobility (termed as mobility), 2) death counts, 3) mean cases, 4) effective reproduction number (R_t_), 5) hospitalization, and 6) mean infections from the Institute for Health Metrics and Evaluations (IHME) from 13^th^ March to 20^th^ August 2020 for 30 Indian states and union territories (UTs). National and subnational mid-year population data were extracted from the Census-based Population Council projections for 2020. The first lockdown phase (LP1) was from 25^th^ March to 14^th^ April 2020 (21 days), the second phase (LP2) was from 15^th^ April to 3^rd^ May 2020 (19 days), the third phase (LP3) was from 4^th^ to 17^th^ May 2020 (14 days), and the fourth lockdown phase (LP4) was from 18^th^ to 31^st^ May 2020 (14 days). The first unlock phase (UP1) was from 1^st^ to 30^th^ June 2020 (30 days) while the second unlock phase (UP2) was from 1^st^ to 31^st^ July 2020 (31 days) ^21^. Our study duration lasts 159 days from March 14^th^ until August 19^th^ 2020, which extends beyond the unlock intervention periods to analyze the after-effects of the interventions.

### Data Variables

Our analysis focuses on six COVID-19 pandemic outcomes, 1) case growth C(t), 2) death count D(t), 3) effective reproductive number (R_t_), 4) composite mobility M(t), 5) infection growth, and 6) hospitalization. First, case growth is calculated as

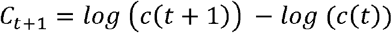

where *c*(*t*) represents the average number of cases observed on day *t*.

To understand the trend in these cases, we converted them to the log scale, and calculating the rolling difference for consecutive days allows us to evaluate the growth^33^. Similar to case growth, we also defined infection growth as a rolling difference in the infections for consecutive days on a log scale. Next, we considered the death counts in each region as another outcome. We used the normalized composite mobility index, *M*_*t*_, which accounts for the change in mobility and ranges between [-100,0] during our study period^34^, where 0 indicates typical mobility or the baseline.

### Data analysis

Two statistical methods used for assessing the impact of the interventions across the four outcomes of interest are described below:

#### 1) Interrupted time series regression (ITR)

For performing the ITR analysis, we required information about when the interventions were implemented, and the time elapsed since the previous intervention for each time point in our data. To do this we represented 4 lockdown and 2 unlock phases using indicator variables. For example, we set the phase indicator for the first lockdown phase as 1 for all the dates between 25th March 2020 to 14th April 2020 which comprises the first lockdown period, and set is 0 for all the remaining dates for the mobility indicator. However, to account for the lags in reporting and transmission for other outcomes, additional days were incorporated; namely +7 for R_t_, +10 for *C*_*t*_, and +13 for *D*_*t*_, +14 for hospitalizations, and +7 for infection rate. Thus, phase 1 *R*_*t*_ for lasted from April 1 to April 21 2020, for *C*_*t*_ lasted from April 4 until April 24 2020, and for *D*_*t*_ lasted from April 7 until April 27 2020. We follow a similar procedure to set indicators for the other lockdown and unlock phases.

To identify their variance at the sub-national level for the effect of the interventions, we ran the following models for each outcome and phase and choose the one that best explains the data for the specific outcome and phase:

1. Fixed effects model: *log* (*Y*_*t*_) = *β*_0_ + *eβ*_1_*N*_0_ + *β*_2_ *P*_*i*_ + *β*_3_ *N*_1_ + *β*_4_. *log* (*population*)
2. Random intercept model: *log* (*Y*_*t*_) = *β*_0_ + *β*_1_*N*_0_ + *β*_2_*P*_*i*_ + *β*_3_*N*_1_ + *β*_4_. *log* (*population*) + (1|*State*)
3. Random slopes model: *log* (*Y*_*t*_) = *β*_0_ + *β*_1_*N*_0_ + *β*_2_*P*_*i*_ + *β*_3_*N*_1_ + *β*_4_. *log* (*population*) + (0 + *N*_0_ + *log* (*population*)|*State*)
4. Mixed effects model: *log* (*Y*_*t*_) *= β*_0_ + *β*_1_*N*_0_ + *β*_2_*P*_*i*_ + *β*_3_*N*_1_ + *β*_4_. *log* (*population*) + (1 + *N*_0_ + *log* (*population*)|*State*)

Where:

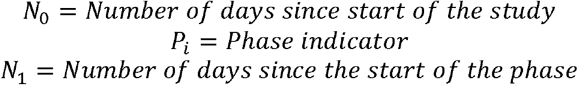

and Y represents the six outcomes of interest. Finally, we obtained 36 models for the 6 outcomes and 6 phases. In the multi-level models, phase indicator variables and the number of days since the last intervention phase were used as fixed effects while the states and UTs. For each model, *β*_0_ represents the baseline level of the outcome at t=0, *β*_1_ represents the change in the outcome per day pre-intervention, *β*_2_ represents the change in the level of the outcome immediately post-intervention, and *β*_3_, is our primary parameter of interest, which represents the difference in the slope post-intervention compared to pre-intervention period.

We determine the model fit for different models using AIC values.

#### 2) Bayesian causal impact analysis

As we observed variance in the estimates at the sub-national level with the ITR analysis, Bayesian causal impact analysis is performed for each state. This analysis centers around a Bayesian Structural time series model aimed to estimate the effect of an intervention. The effect is assessed by comparing the observed outcome values after an intervention is performed and the baseline values; the Bayesian structural time series model predicts the outcome for the post-intervention period in the absence of the intervention. Thus, for the causal impact analysis, we have the outcome observed during our study time as well as the dates on which interventions were performed as the input. The causal impact model builds on this data to forecast the values had the intervention not taken place, i.e., the counterfactual values in the post-intervention period. Bayesian causal impact analysis thus allows estimating the causal effect by comparing the observed and counterfactual values whereas ITR helps in determining the effect by comparing a change in the coefficients of the predictors before and after an intervention, although both aim to answer causal questions.

We implemented six models for each outcome and separate models for each region (state/UT). Thus, our primary analysis consisted 6 * 6 * (30 (number of states and union territories at a subnational level) + 1(aggregate data at the national level), i.e., a total of 1080 models. The dates of intervention for the different outcomes and phases are similar to the processing done for ITR which account for lag for all the outcomes. As most of the potential covariates that could be included in the analysis and which are associated with the outcome such as mobility, weather conditions, temperature, and pollution were impacted by the lockdown and unlock interventions, i.e., they were not independent of the interventions, we did not include any covariates in the causal impact analysis. We also did not incorporate seasonality features as the assessment depicted a lack of significant seasonality. Thus, default parameters were used with dynamic regression and seasonality set to False. The model is described below:

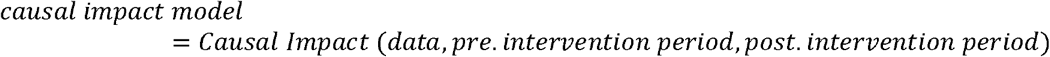

We obtain the relative and absolute effect of each intervention for each outcome along with the p-values using the causal impact analysis. To understand the effect of each intervention, we flip (negate) the effects of the unlock phases since the nature of the unlock intervention is opposite to that of the lockdown intervention. Furthermore, we normalize the effects for all states and UTs between 0 and 1 across all outcomes to compare across outcomes. We do this independently for the 4 lockdown phases and the 2 unlock phases. Accordingly, **Figure 2** represents the causal effects for all states and outcomes normalized for the 4 lockdown phases in the left panel and the normalized causal effects across all states and outcomes for the 2 unlock phases in the right panel. We use the geopandas package in Python to visualize the intervention effects across outcomes and phases. We also conducted sensitivity analyses (+/-3 days) around the intervention periods by considering additional lag periods (different across outcomes) around the intervention phase dates but found that the results did not vary based on sensitivity. The statistical analysis is carried out in R version 4.0.3, epinow2, CausalImpact, lme4 packages for the main analysis. Data processing and plots are generated using Python version 3.5. The code is made publicly available at https://github.com/asarforindia/India-lockdown-impact.

## Data Availability

All data produced are available online at https://github.com/asarforindia/India-lockdown-impact

## Acknowledgements

None

## SUPPLEMENTARY MATERIAL

**Table S1:** **Absolute causal effect of intervention [with 95% credible interval] as computed using causal impact analysis for different outcomes. Rows represent different intervention phases and columns represent the outcomes with sensitivity to the intervention: date of intervention - the relevant time window.**

|  | Outcome (95% Credible Interval) |  |  |  |
| --- | --- | --- | --- | --- |
| Phase | *Case growth* | *Death rate* | $R_t$ | *Mobility* |
| *Lockdown phase 1* | -0.285<br>[-0.794, 0.197] | 0.001<br>[0.001, 0.001] | -0.243<br>[-0.307, -0.179] | -23.502<br>[-41.087, -6.654] |
| *Lockdown phase 2* | -0.041<br>[-0.318, 0.245] | 0.002<br>[0.001, 0.002] | 0.379<br>[0.275, 0.480] | 41.340<br>[27.079, 54.529] |
| *Lockdown phase 3* | -0.022<br>[-0.219, 0.191] | 0.002<br>[0.001, 0.003] | 0.221<br>[0.127, 0.326] | 28.237<br>[17.180, 38.594] |
| *Lockdown phase 4* | -0.014<br>[-0.198, 0.170] | 0.007<br>(0.006, 0.007) | 0.200<br>[0.108, 0.288] | 25.705<br>[15.566, 36.149] |
| *Unlock phase 1* | 0.009<br>[-0.162, 0.149] | 0.006<br>[0.005, 0.007] | 0.180<br>[0.063, 0.297] | 26.835<br>[17.944, 35.316] |
| *Unlock phase 2* | 0.011<br>[-0.114, 0.134] | 0.011<br>[0.009, 0.014] | 0.092<br>[-0.027, 0.226] | 0.314<br>[-7.754, 8.332] |

**Table S2:**
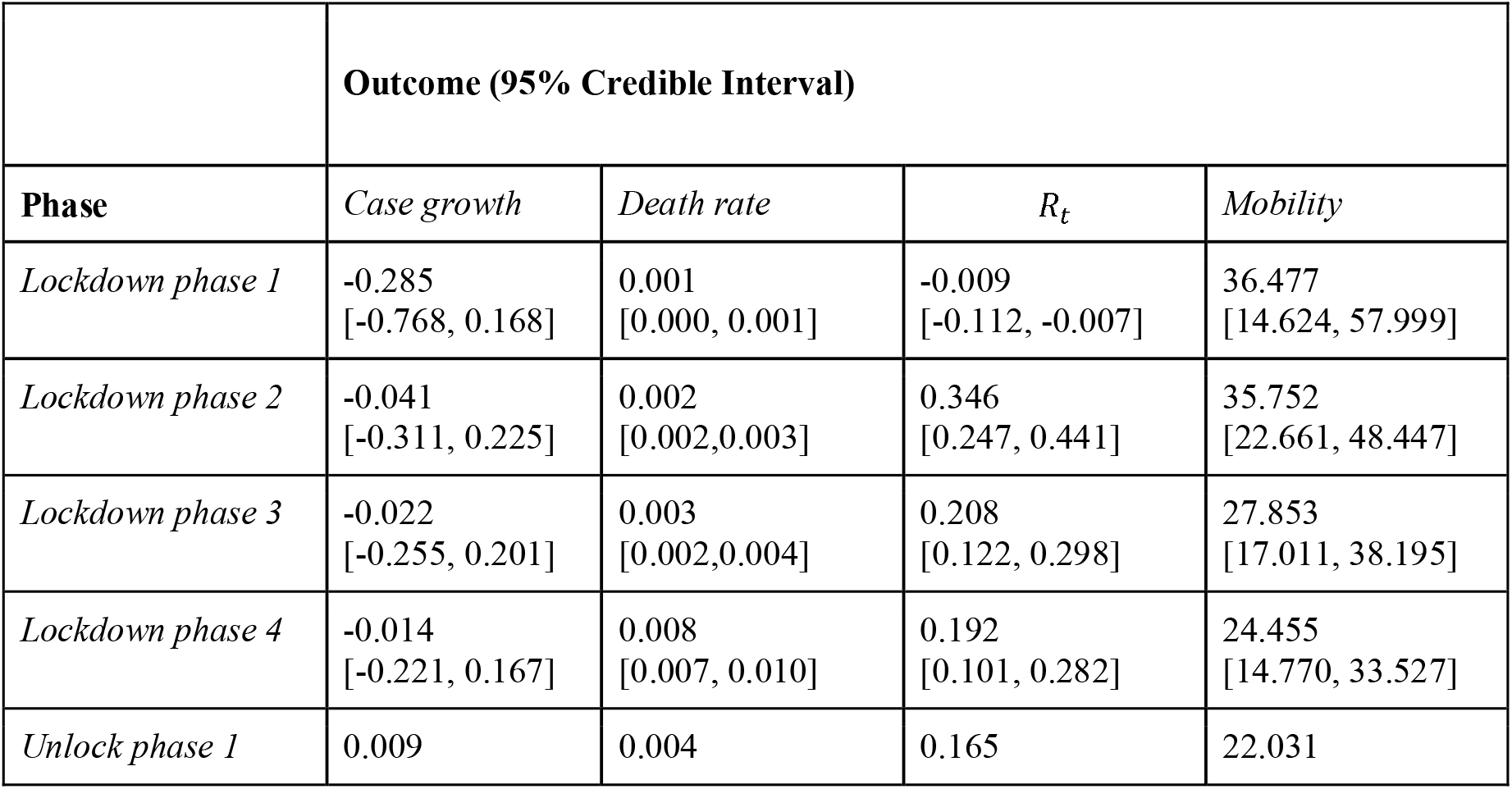
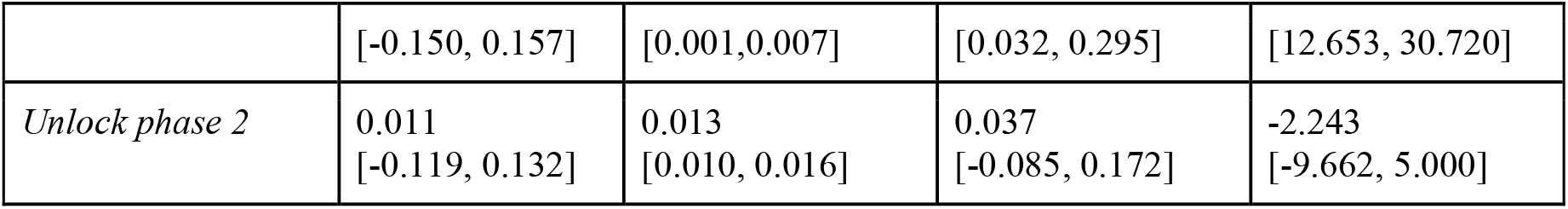
**Absolute causal effect of intervention [with 95% credible interval] as computed using causal impact analysis for different outcomes. Rows represent different intervention phases and columns represent the outcomes with sensitivity to the intervention: date of intervention + the relevant time window.**

**Table S4:** Estimates for case growth in lockdown phase 1 using ITR. (case_growth∼days0+days1+phase_indi_0+log_sum_pop+(0+days0+log_sum_pop|state), AIC: - 5068.7844)

|  | Estimate | 2.5_ci | 97.5_ci | SE | DF | T-stat | P-val | Sig |
| --- | --- | --- | --- | --- | --- | --- | --- | --- |
| (Intercept) | -0.0654592 | -0.1161346 | -0.0147838 | 0.02585529 | 754.563762 | -2.5317525 | 0.01155127 | * |
| days0 | 0.00018214 | -0.0010385 | 0.00140278 | 0.00062279 | 4769.51854 | 0.29246259 | 0.76994568 |  |
| days1 | -0.0006662 | -0.0019512 | 0.00061878 | 0.00065562 | 4896.59044 | -1.0161572 | 0.30960473 |  |
| phase_indi_0 | -0.0531014 | -0.0674616 | -0.0387412 | 0.00732677 | 4896.59038 | -7.2475848 | 4.91E-13 | *** |
| log_sum_pop | 0.00807132 | 0.00529021 | 0.01085243 | 0.00141896 | 696.657568 | 5.68819663 | 1.89E-08 | *** |

**(a) Fixed effects**
|  | Name | Var | Std |
| --- | --- | --- | --- |
| state | days0 | 1.73E-07 | 0.00041643 |
| state | log_sum_pop | 2.62E-06 | 0.00161904 |
| Residual |  | 0.02052475 | 0.14326461 |

**Table S5:** Estimates for case growth in lockdown phase 2 using ITR.

|  | Estimate | 2.5_ci | 97.5_ci | SE | DF | T-stat | P-val | Sig |
| --- | --- | --- | --- | --- | --- | --- | --- | --- |
| (Intercept) | -0.0518497 | -0.102708 | -0.0009914 | 0.02594858 | 735.746445 | -1.9981711 | 0.04606573 | * |
| days0 | -0.0014368 | -0.0026024 | -0.0002712 | 0.00059469 | 4731.93518 | -2.4160559 | 0.01572728 | * |
| days1 | 0.00118378 | -3.01E-05 | 0.00239763 | 0.00061932 | 4896.52927 | 1.91141937 | 0.05600903 | . |
| phase_indi_1 | -0.0081309 | -0.0213975 | 0.0051356 | 0.00676877 | 4896.52932 | -1.2012442 | 0.22971457 |  |
| log_sum_pop | 0.00807943 | 0.00528392 | 0.01087494 | 0.00142631 | 680.717113 | 5.6645789 | 2.18E-08 | *** |

**(a) Fixed effects**
|  | Name | Var | Std |
| --- | --- | --- | --- |
| state | days0 | 1.72E-07 | 0.00041427 |
| state | log_sum_pop | 2.58E-06 | 0.00160662 |
| Residual |  | 0.02073936 | 0.14401168 |

**Table S6:** Estimates for case growth in lockdown phase 3 using ITR.

|  | Estimate | 2.5_ci | 97.5_ci | SE | DF | T-stat | P-val | Sig |
| --- | --- | --- | --- | --- | --- | --- | --- | --- |
| (Intercept) | -0.0478624 | -0.0986769 | 0.00295206 | 0.02592624 | 710.249771 | -1.8461003 | 0.06529358 | . |
| days0 | -0.0019942 | -0.0031309 | -0.0008574 | 0.00058 | 4712.70431 | -3.4382702 | 0.00059051 | *** |
| days1 | 0.00179609 | 0.00061756 | 0.00297462 | 0.0006013 | 4896.43539 | 2.98700162 | 0.00283125 | ** |
| phase_indi_2 | 0.02945581 | 0.0150448 | 0.04386681 | 0.00735269 | 4896.43546 | 4.00612702 | 6.26E-05 | *** |
| log_sum_pop | 0.00812338 | 0.0053282 | 0.01091856 | 0.00142614 | 654.505871 | 5.69607358 | 1.85E-08 | *** |

**(a) Fixed effects**
|  | Name | Var | Std |
| --- | --- | --- | --- |
| state | days0 | 1.70E-07 | 0.00041242 |
| state | log_sum_pop | 2.53E-06 | 0.00159091 |
| Residual |  | 0.02067823 | 0.14379926 |

**Table S7:** Estimates for case growth in lockdown phase 4 using ITR.

|  | Estimate | 2.5_ci | 97.5_ci | SE | DF | T-stat | P-val | Sig |
| --- | --- | --- | --- | --- | --- | --- | --- | --- |
| (Intercept) | -0.048718 | -0.0994925 | 0.00205641 | 0.0259058 | 729.833117 | -1.8805838 | 0.06042652 | . |
| days0 | -0.0018115 | -0.0029413 | -0.0006816 | 0.00057646 | 4709.65825 | -3.1424239 | 0.00168595 | ** |
| days1 | 0.00158746 | 0.00041757 | 0.00275735 | 0.00059689 | 4896.34237 | 2.65953855 | 0.00785026 | ** |
| phase_indi_3 | 0.02159342 | 0.00732947 | 0.03585737 | 0.00727766 | 4896.34228 | 2.96708233 | 0.00302095 | ** |
| log_sum_pop | 0.00808139 | 0.00528819 | 0.01087459 | 0.00142513 | 676.605502 | 5.67063793 | 2.11E-08 | *** |

**(a) Fixed effects**
|  | Name | Var | Std |
| --- | --- | --- | --- |
| state | days0 | 1.69E-07 | 0.00041134 |
| state | log_sum_pop | 2.54E-06 | 0.00159332 |
| Residual |  | 0.02070952 | 0.14390802 |

**Table S8:** Estimates for case growth in unlock phase 1 using ITR.

|  | Estimate | 2.5_ci | 97.5_ci | SE | DF | T-stat | P-val | Sig |
| --- | --- | --- | --- | --- | --- | --- | --- | --- |
| (Intercept) | -0.0499486 | -0.1006773 | 0.00078006 | 0.02588245 | 756.477404 | -1.9298255 | 0.05400207 | . |
| days0 | -0.0016096 | -0.0027356 | -0.0004836 | 0.0005745 | 4655.19959 | -2.8017424 | 0.00510374 | ** |
| days1 | 0.0013855 | 0.00022115 | 0.00254984 | 0.00059407 | 4897.07163 | 2.33222708 | 0.01972907 | * |
| phase_indi_4 | -0.0029923 | -0.0136312 | 0.00764656 | 0.0054281 | 4897.07171 | -0.5512655 | 0.58147682 |  |
| log_sum_pop | 0.00805031 | 0.00525533 | 0.01084529 | 0.00142604 | 708.999794 | 5.6452311 | 2.39E-08 | *** |

**(a) Fixed effects**
|  | Name | Var | Std |
| --- | --- | --- | --- |
| state | days0 | 1.80E-07 | 0.00042401 |
| state | log_sum_pop | 2.74E-06 | 0.00165547 |
| Residual |  | 0.02074039 | 0.14401525 |

**Table S9:** Estimates for case growth in unlock phase 2 using ITR.

|  | Estimate | 2.5_ci | 97.5_ci | SE | DF | T-stat | P-val | Sig |
| --- | --- | --- | --- | --- | --- | --- | --- | --- |
| (Intercept) | -0.0514642 | -0.1022666 | -0.0006617 | 0.02592009 | 699.747258 | -1.9854942 | 0.04747942 | * |
| days0 | -0.0015112 | -0.0026493 | -0.0003731 | 0.00058066 | 4601.43315 | -2.6025811 | 0.00928217 | ** |
| days1 | 0.00123056 | 4.08E-05 | 0.00242034 | 0.00060704 | 4897.86543 | 2.02714665 | 0.04270147 | * |
| phase_indi_5 | 0.00850321 | -0.0043425 | 0.02134893 | 0.00655406 | 4897.86542 | 1.29739475 | 0.1945565 |  |
| log_sum_pop | 0.00808565 | 0.00528241 | 0.01088888 | 0.00143025 | 636.708517 | 5.65331908 | 2.38E-08 | *** |

**(a) Fixed effects**
|  | Name | Var | Std |
| --- | --- | --- | --- |
| state | days0 | 1.93E-07 | 0.00043909 |
| state | log_sum_pop | 2.95E-06 | 0.00171672 |
| Residual |  | 0.02072832 | 0.14397332 |

**Table S10:**
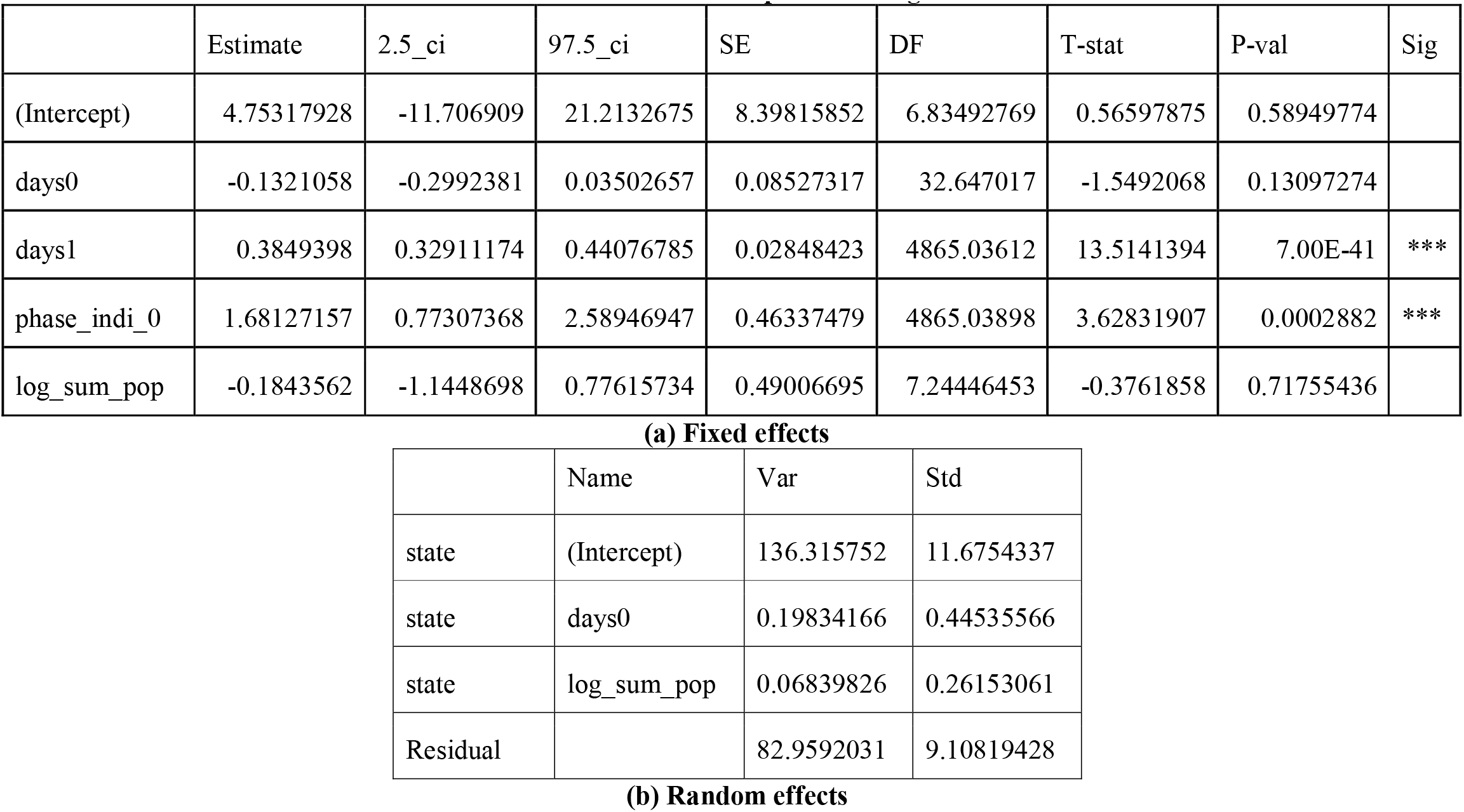
Estimates for death count in lockdown phase 1 using ITR.

|  | Estimate | 2.5_ci | 97.5_ci | SE | DF | T-stat | P-val | Sig |
| --- | --- | --- | --- | --- | --- | --- | --- | --- |
| (Intercept) | 4.75317928 | -11.706909 | 21.2132675 | 8.39815852 | 6.83492769 | 0.56597875 | 0.58949774 |  |
| days0 | -0.1321058 | -0.2992381 | 0.03502657 | 0.08527317 | 32.647017 | -1.5492068 | 0.13097274 |  |
| days1 | 0.3849398 | 0.32911174 | 0.44076785 | 0.02848423 | 4865.03612 | 13.5141394 | 7.00E-41 | *** |
| phase_indi_0 | 1.68127157 | 0.77307368 | 2.58946947 | 0.46337479 | 4865.03898 | 3.62831907 | 0.0002882 | *** |
| log_sum_pop | -0.1843562 | -1.1448698 | 0.77615734 | 0.49006695 | 7.24446453 | -0.3761858 | 0.71755436 |  |

**(a) Fixed effects**
|  | Name | Var | Std |
| --- | --- | --- | --- |
| state | (Intercept) | 136.315752 | 11.6754337 |
| state | days0 | 0.19834166 | 0.44535566 |
| state | log_sum_pop | 0.06839826 | 0.26153061 |
| Residual |  | 82.9592031 | 9.10819428 |

**Table S11:** Estimates for death count in lockdown phase 2 using ITR.

**(b) Random effects****Table S11: Estimates for death count in lockdown phase 2 using ITR**
|  | Estimate | 2.5_ci | 97.5_ci | SE | DF | T-stat | P-val | Sig |
| --- | --- | --- | --- | --- | --- | --- | --- | --- |
| (Intercept) | -1.3848926 | -13.02219 | 10.2524049 | 5.93750579 | 6.16838833 | -0.2332448 | 0.82311937 |  |
| days0 | -0.0678661 | -0.2335677 | 0.09783542 | 0.08454316 | 34.6311021 | -0.8027395 | 0.42760032 |  |
| days1 | 0.30834641 | 0.25678786 | 0.35990497 | 0.02630587 | 4871.83859 | 11.7215835 | 2.59E-31 | *** |
| phase_indi_1 | -1.4458886 | -2.2800238 | -0.6117534 | 0.42558701 | 4871.83866 | -3.3973982 | 0.00068575 | *** |
| log_sum_pop | 0.13931836 | -0.5520184 | 0.83065517 | 0.35272934 | 30.3868206 | 0.39497243 | 0.69562221 |  |

**(a) Fixed effects**
|  | Name | Var | Std |
| --- | --- | --- | --- |
| state | (Intercept) | 696.05105 | 26.3827794 |
| state | days0 | 0.197003 | 0.44385019 |
| state | log_sum_pop | 2.34870883 | 1.53254978 |
| Residual |  | 82.9296254 | 9.10657045 |

**Table S12:** Estimates for death count in lockdown phase 3 using ITR.

|  | Estimate | 2.5_ci | 97.5_ci | SE | DF | T-stat | P-val | Sig |
| --- | --- | --- | --- | --- | --- | --- | --- | --- |
| (Intercept) | 4.03844242 | -6.165232 | 14.2421168 | 5.20605198 | 38.998454 | 0.77572073 | 0.44259285 |  |
| days0 | -0.0781487 | -0.2442526 | 0.08795528 | 0.08474847 | 33.9712868 | -0.9221248 | 0.36296651 |  |
| days1 | 0.32112125 | 0.27129736 | 0.37094514 | 0.02542082 | 4873.16 | 12.6322153 | 5.12E-36 | *** |
| phase_indi_2 | -1.5603908 | -2.4713925 | -0.649389 | 0.46480534 | 4873.16005 | -3.3570844 | 0.00079372 | *** |
| log_sum_pop | -0.1564782 | -0.8349572 | 0.52200087 | 0.34616913 | 29.8930503 | -0.4520281 | 0.65451184 |  |

**(a) Fixed effects**
|  | Name | Var | Std |
| --- | --- | --- | --- |
| state | (Intercept) | 422.524587 | 20.5554029 |
| state | days0 | 0.19900536 | 0.44610017 |
| state | log_sum_pop | 2.09936522 | 1.44891864 |
| Residual |  | 82.9591072 | 9.10818902 |

**Table S13:**
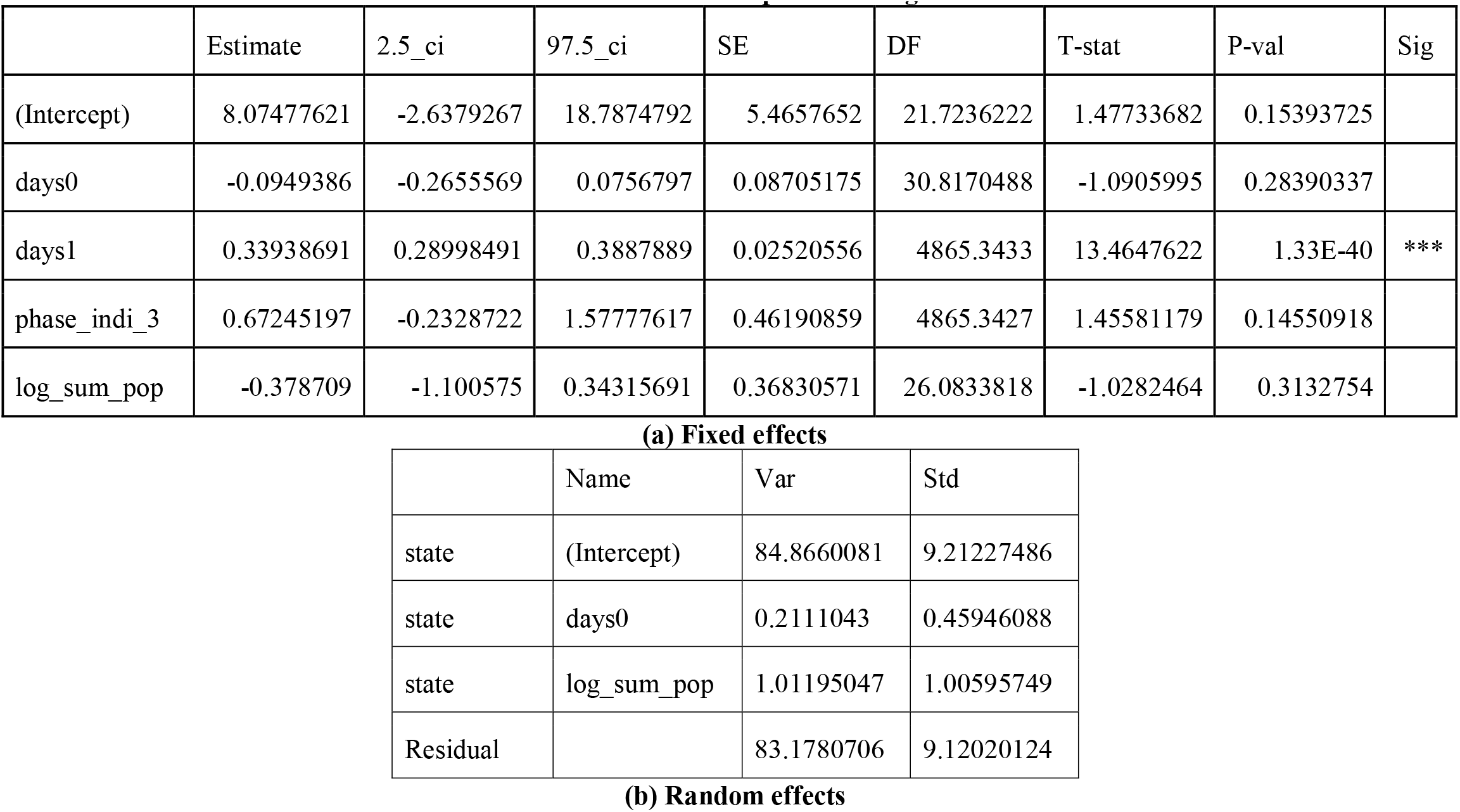
Estimates for death count in lockdown phase 4 using ITR.

**(b) Random effects****Table S13: Estimates for death count in lockdown phase 4 using ITR**
|  | Estimate | 2.5_ci | 97.5_ci | SE | DF | T-stat | P-val | Sig |
| --- | --- | --- | --- | --- | --- | --- | --- | --- |
| (Intercept) | 8.07477621 | -2.6379267 | 18.7874792 | 5.4657652 | 21.7236222 | 1.47733682 | 0.15393725 |  |
| days0 | -0.0949386 | -0.2655569 | 0.0756797 | 0.08705175 | 30.8170488 | -1.0905995 | 0.28390337 |  |
| days1 | 0.33938691 | 0.28998491 | 0.3887889 | 0.02520556 | 4865.3433 | 13.4647622 | 1.33E-40 | *** |
| phase_indi_3 | 0.67245197 | -0.2328722 | 1.57777617 | 0.46190859 | 4865.3427 | 1.45581179 | 0.14550918 |  |
| log_sum_pop | -0.378709 | -1.100575 | 0.34315691 | 0.36830571 | 26.0833818 | -1.0282464 | 0.3132754 |  |

**(a) Fixed effects**
|  | Name | Var | Std |
| --- | --- | --- | --- |
| state | (Intercept) | 84.8660081 | 9.21227486 |
| state | days0 | 0.2111043 | 0.45946088 |
| state | log_sum_pop | 1.01195047 | 1.00595749 |
| Residual |  | 83.1780706 | 9.12020124 |

**Table S14:** Estimates for death count in unlock phase 1 using ITR.

|  | Estimate | 2.5_ci | 97.5_ci | SE | DF | T-stat | P-val | Sig |
| --- | --- | --- | --- | --- | --- | --- | --- | --- |
| (Intercept) | 4.39079676 | -5.3134969 | 14.0950904 | 4.9512612 | 49.4797884 | 0.8868037 | 0.37947474 |  |
| days0 | -0.0873006 | -0.2541654 | 0.07956429 | 0.08513669 | 33.126512 | -1.0254165 | 0.3125967 |  |
| days1 | 0.33929376 | 0.29037916 | 0.38820836 | 0.02495689 | 4870.10146 | 13.5951955 | 2.41E-41 | *** |
| phase_indi_4 | -2.492173 | -3.1811619 | -1.8031841 | 0.35153141 | 4870.10144 | -7.0894747 | 1.54E-12 | *** |
| log_sum_pop | -0.1715427 | -0.8170701 | 0.47398466 | 0.32935675 | 39.3826657 | -0.5208416 | 0.60539343 |  |

**(a) Fixed effects**
|  | Name | Var | Std |
| --- | --- | --- | --- |
| state | (Intercept) | 330.827382 | 18.1886608 |
| state | days0 | 0.20155387 | 0.44894751 |
| state | log_sum_pop | 1.70268668 | 1.30487037 |
| Residual |  | 82.334766 | 9.07385067 |

**Table S15:** Estimates for death count in unlock phase 2 using ITR.

**(b) Random effects****Table S15: Estimates for death count in unlock phase 2 using ITR**
|  | Estimate | 2.5_ci | 97.5_ci | SE | DF | T-stat | P-val | Sig |
| --- | --- | --- | --- | --- | --- | --- | --- | --- |
| (Intercept) | 6.28234217 | -3.0342381 | 15.5989224 | 4.75344462 | 76.1754221 | 1.32163992 | 0.19024367 |  |
| days0 | -0.0456543 | -0.2123524 | 0.12104385 | 0.08505163 | 33.9794744 | -0.5367831 | 0.59491498 |  |
| days1 | 0.26302798 | 0.21063848 | 0.31541748 | 0.02672983 | 4868.2197 | 9.84024229 | 1.23E-22 | *** |
| phase_indi_5 | 3.58294864 | 2.65402344 | 4.51187385 | 0.47395014 | 4868.21986 | 7.55975862 | 4.80E-14 | *** |
| log_sum_pop | -0.30645 | -0.9519356 | 0.33903563 | 0.32933546 | 45.856931 | -0.9305102 | 0.35697992 |  |

**(a) Fixed effects**
|  | Name | Var | Std |
| --- | --- | --- | --- |
| state | (Intercept) | 215.060095 | 14.6649274 |
| state | days0 | 0.20005034 | 0.44726988 |
| state | log_sum_pop | 1.48149456 | 1.21716661 |
| Residual |  | 82.2602492 | 9.06974361 |

**Table S16:**
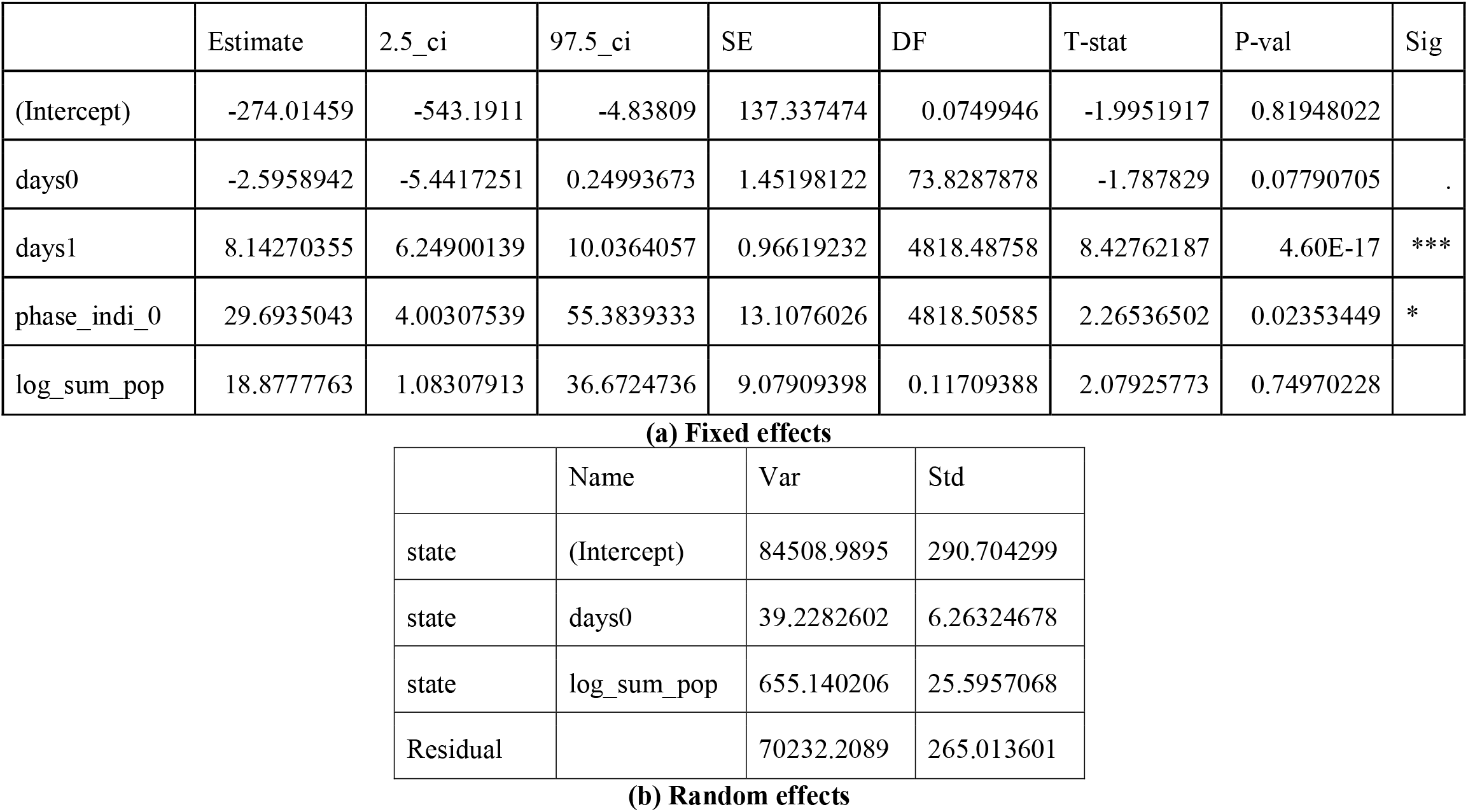
Estimates for hospitalization in lockdown phase 1 using ITR.

**Table S17:** Estimates for hospitalization in lockdown phase 2 using ITR.

**(b) Random effects****Table S17: Estimates for hospitalization in lockdown phase 2 using ITR**
|  | Estimate | 2.5_ci | 97.5_ci | SE | DF | T-stat | P-val | Sig |
| --- | --- | --- | --- | --- | --- | --- | --- | --- |
| (Intercept) | -289.3045 | -553.64839 | -24.960612 | 134.871809 | 1.52073959 | -2.1450331 | 0.20378491 |  |
| days0 | -1.135032 | -3.9244538 | 1.65438976 | 1.42320052 | 69.5892038 | -0.7975208 | 0.42786158 |  |
| days1 | 6.4576034 | 4.67767209 | 8.23753471 | 0.90814491 | 4865.63427 | 7.11076322 | 1.32E-12 | *** |
| phase_indi_1 | -37.07525 | -61.089592 | -13.060908 | 12.2524406 | 4865.6341 | -3.0259482 | 0.00249151 | ** |
| log_sum_pop | 18.8821766 | 1.39092068 | 36.3734326 | 8.92427416 | 2.28798845 | 2.11582211 | 0.15235964 |  |

**(a) Fixed effects**
|  | Name | Var | Std |
| --- | --- | --- | --- |
| state | (Intercept) | 92073.0509 | 303.435415 |
| state | days0 | 39.0657249 | 6.25025799 |
| state | log_sum_pop | 672.606963 | 25.9346672 |
| Residual |  | 70178.6644 | 264.91256 |

**Table S18:**
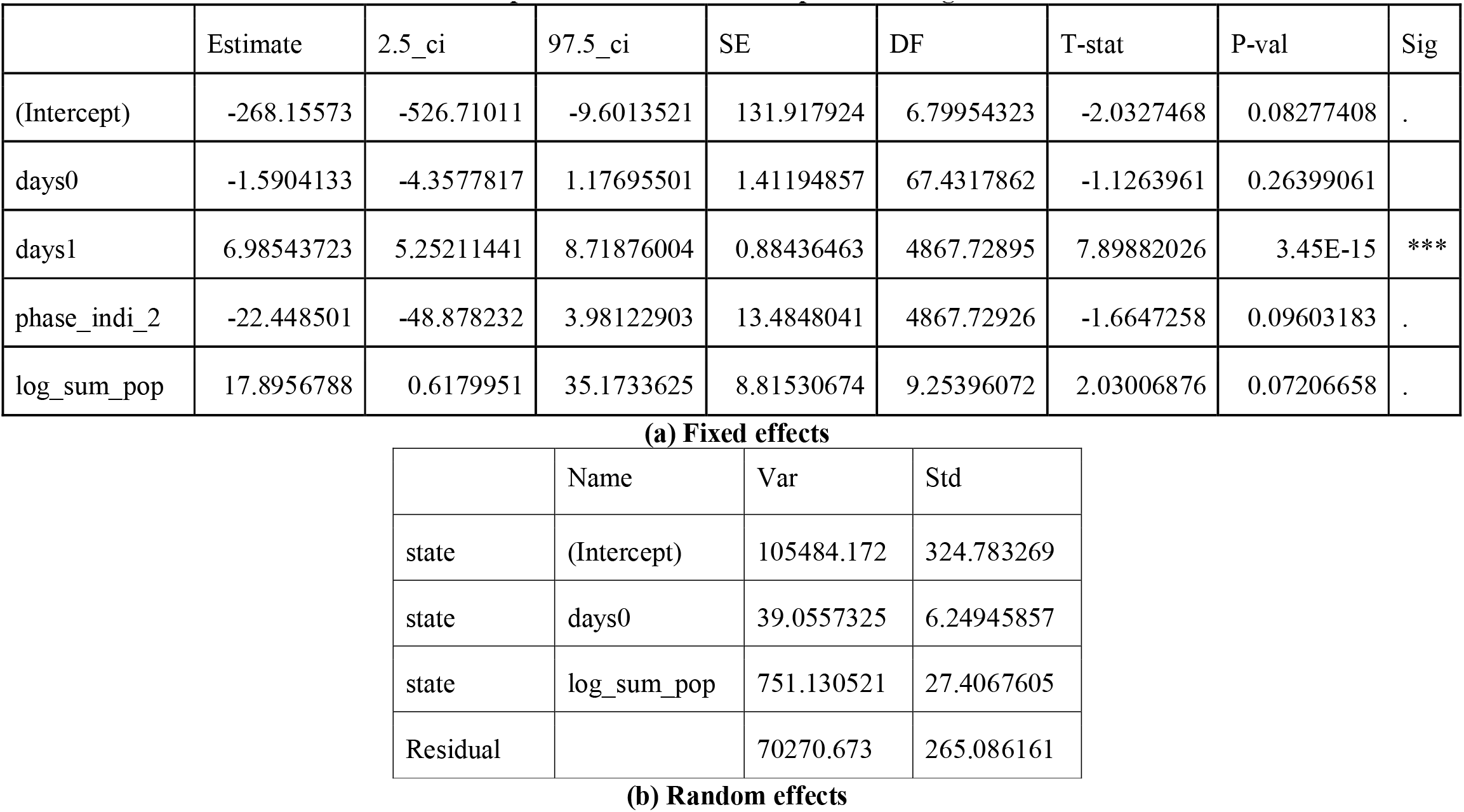
Estimates for hospitalization in lockdown phase 3 using ITR.

**(b) Random effects****Table S18: Estimates for hospitalization in lockdown phase 3 using ITR**
|  | Estimate | 2.5_ci | 97.5_ci | SE | DF | T-stat | P-val | Sig |
| --- | --- | --- | --- | --- | --- | --- | --- | --- |
| (Intercept) | -268.15573 | -526.71011 | -9.6013521 | 131.917924 | 6.79954323 | -2.0327468 | 0.08277408 | . |
| days0 | -1.5904133 | -4.3577817 | 1.17695501 | 1.41194857 | 67.4317862 | -1.1263961 | 0.26399061 |  |
| days1 | 6.98543723 | 5.25211441 | 8.71876004 | 0.88436463 | 4867.72895 | 7.89882026 | 3.45E-15 | *** |
| phase_indi_2 | -22.448501 | -48.878232 | 3.98122903 | 13.4848041 | 4867.72926 | -1.6647258 | 0.09603183 | . |
| log_sum_pop | 17.8956788 | 0.6179951 | 35.1733625 | 8.81530674 | 9.25396072 | 2.03006876 | 0.07206658 | . |

**(a) Fixed effects**
|  | Name | Var | Std |
| --- | --- | --- | --- |
| state | (Intercept) | 105484.172 | 324.783269 |
| state | days0 | 39.0557325 | 6.24945857 |
| state | log_sum_pop | 751.130521 | 27.4067605 |
| Residual |  | 70270.673 | 265.086161 |

**Table S19:** Estimates for hospitalization in lockdown phase 4 using ITR.

**(b) Random effects****Table S19: Estimates for hospitalization in lockdown phase 4 using ITR**
|  | Estimate | 2.5_ci | 97.5_ci | SE | DF | T-stat | P-val | Sig |
| --- | --- | --- | --- | --- | --- | --- | --- | --- |
| (Intercept) | -260.21619 | -522.59522 | 2.16283878 | 133.869313 | 3.57271498 | -1.9438076 | 0.13228499 |  |
| days0 | -1.6043653 | -4.3595449 | 1.15081435 | 1.40572972 | 67.6849946 | -1.1413042 | 0.25776526 |  |
| days1 | 7.01936716 | 5.30010458 | 8.73862974 | 0.87719091 | 4867.15448 | 8.0020975 | 1.52E-15 | *** |
| phase_indi_3 | -33.047976 | -59.322421 | -6.7735311 | 13.4055754 | 4867.15516 | -2.4652411 | 0.01372637 | * |
| log_sum_pop | 17.4864358 | -0.0388756 | 35.0117471 | 8.94164968 | 5.24300054 | 1.95561629 | 0.10522709 |  |

**(a) Fixed effects**
|  | Name | Var | Std |
| --- | --- | --- | --- |
| state | (Intercept) | 98228.9977 | 313.415057 |
| state | days0 | 38.8211858 | 6.23066496 |
| state | log_sum_pop | 733.522358 | 27.0836179 |
| Residual |  | 70222.9283 | 264.996091 |

**Table S20:** Estimates for hospitalization in unlock phase 1 using ITR.

|  | Estimate | 2.5_ci | 97.5_ci | SE | DF | T-stat | P-val | Sig |
| --- | --- | --- | --- | --- | --- | --- | --- | --- |
| (Intercept) | -615.1556 | -1035.6338 | -194.6774 | 214.533636 | 3.24520769 | -2.8674086 | 0.05845559 | . |
| days0 | -1.7313844 | -4.4863088 | 1.02353989 | 1.40559947 | 56.1065654 | -1.2317765 | 0.22317028 |  |
| days1 | 7.31835641 | 5.61015713 | 9.02655568 | 0.87154626 | 4868.07059 | 8.39697989 | 5.94E-17 | *** |
| phase_indi_4 | -57.045984 | -77.111205 | -36.980762 | 10.237546 | 4868.0713 | -5.5722322 | 2.65E-08 | *** |
| log_sum_pop | 38.0056145 | 13.4391072 | 62.5721218 | 12.5341626 | 3.80534634 | 3.03216224 | 0.0412908 | * |

**(a) Fixed effects**
|  | Name | Var | Std |
| --- | --- | --- | --- |
| state | (Intercept) | 84806.0883 | 291.214849 |
| state | days0 | 39.1035171 | 6.25328051 |
| state | log_sum_pop | 51.4082735 | 7.16995631 |
| Residual |  | 69823.8652 | 264.242058 |

**Table S21:** Estimates for hospitalization in unlock phase 2 using ITR.

|  | Estimate | 2.5_ci | 97.5_ci | SE | DF | T-stat | P-val | Sig |
| --- | --- | --- | --- | --- | --- | --- | --- | --- |
| (Intercept) | -181.85666 | -447.8875 | 84.1741756 | 135.732514 | 41.5642102 | -1.3398165 | 0.18758454 |  |
| days0 | -0.0107264 | -2.7596351 | 2.73818233 | 1.40253021 | 72.3613334 | -0.0076479 | 0.99391899 |  |
| days1 | 4.56453995 | 2.7632714 | 6.36580851 | 0.91903146 | 4871.31962 | 4.96668522 | 7.04E-07 | *** |
| phase_indi_5 | 118.362864 | 91.7282342 | 144.997494 | 13.5893466 | 4871.31969 | 8.70997465 | 4.10E-18 | *** |
| log_sum_pop | 11.9055746 | -6.492547 | 30.3036961 | 9.38696919 | 23.9451288 | 1.26830869 | 0.21688071 |  |

**(a) Fixed effects**
|  | Name | Var | Std |
| --- | --- | --- | --- |
| state | (Intercept) | 239235.988 | 489.117561 |
| state | days0 | 37.7681493 | 6.14557965 |
| state | log_sum_pop | 1452.47497 | 38.1113496 |
| Residual |  | 69201.2657 | 263.061334 |

**Table S22:** Estimates for infection growth in lockdown phase 1 using ITR.

|  | Estimate | 2.5_ci | 97.5_ci | SE | DF | T-stat | P-val | Sig |
| --- | --- | --- | --- | --- | --- | --- | --- | --- |
| Intercept | 0.16060766 | 0.12132228 | 0.19989303 | 0.020039 | 4924 | 8.01475349 | 1.33E-15 | *** |
| days0 | -0.0029212 | -0.0041159 | -0.0017264 | 0.00060944 | 4924 | -4.7931966 | 1.69E-06 | *** |
| days1 | 0.00272394 | 0.00147445 | 0.00397342 | 0.00063735 | 4924 | 4.2738487 | 1.96E-05 | *** |
| phase_indi_0 | -0.0192828 | -0.0308041 | -0.0077615 | 0.00587688 | 4924 | -3.2811308 | 0.00104112 | ** |
| log_sum_pop | -0.0037004 | -0.0057528 | -0.0016481 | 0.00104687 | 4924 | -3.5347678 | 0.00041189 | *** |

**Table S23:** Estimates for infection growth in lockdown phase 2 using ITR.

|  | Estimate | 2.5_ci | 97.5_ci | SE | DF | T-stat | P-val | Sig |
| --- | --- | --- | --- | --- | --- | --- | --- | --- |
| Intercept | 0.1730889 | 0.13399927 | 0.21217853 | 0.01993915 | 4924 | 8.68085485 | 0 | *** |
| days0 | -0.0045983 | -0.0057354 | -0.0034612 | 0.00058003 | 4924 | -7.9276666 | 2.66E-15 | *** |
| days1 | 0.00458324 | 0.00340366 | 0.00576282 | 0.00060169 | 4924 | 7.61727375 | 3.09E-14 | *** |
| phase_indi_1 | 0.03294817 | 0.02238504 | 0.0435113 | 0.00538813 | 4924 | 6.11495733 | 1.04E-09 | *** |
| log_sum_pop | -0.0037004 | -0.0057473 | -0.0016536 | 0.00104406 | 4924 | -3.5442914 | 0.00039733 | *** |

**Table S24:** Estimates for infection growth in lockdown phase 3 using ITR.

|  | Estimate | 2.5_ci | 97.5_ci | SE | DF | T-stat | P-val | Sig |
| --- | --- | --- | --- | --- | --- | --- | --- | --- |
| Intercept | 0.16968462 | 0.1305898 | 0.20877944 | 0.0199418 | 4924 | 8.50899242 | 0 | *** |
| days0 | -0.0041419 | -0.0052559 | -0.0030279 | 0.00056825 | 4924 | -7.2888146 | 3.62E-13 | *** |
| days1 | 0.00406977 | 0.00291807 | 0.00522146 | 0.00058747 | 4924 | 6.92764942 | 4.83E-12 | *** |
| phase_indi_2 | 0.03000538 | 0.01851535 | 0.04149541 | 0.00586093 | 4924 | 5.11956143 | 3.18E-07 | *** |
| log_sum_pop | -0.0037004 | -0.0057496 | -0.0016513 | 0.00104524 | 4924 | -3.5402948 | 0.00040338 | *** |

**Table S25:** Estimates for infection growth in lockdown phase 4 using ITR.

|  | Estimate | 2.5_ci | 97.5_ci | SE | DF | T-stat | P-val | Sig |
| --- | --- | --- | --- | --- | --- | --- | --- | --- |
| Intercept | 0.1655651 | 0.12638957 | 0.20474063 | 0.01998297 | 4924 | 8.28531084 | 2.22E-16 | *** |
| days0 | -0.0035854 | -0.004695 | -0.0024757 | 0.00056603 | 4924 | -6.3341794 | 2.60E-10 | *** |
| days1 | 0.00347484 | 0.00232877 | 0.0046209 | 0.0005846 | 4924 | 5.9439917 | 2.97E-09 | *** |
| phase_indi_3 | -0.0101807 | -0.0215594 | 0.00119804 | 0.00580415 | 4924 | -1.7540346 | 0.07948678 | . |
| log_sum_pop | -0.0037004 | -0.0057544 | -0.0016465 | 0.00104769 | 4924 | -3.5320129 | 0.00041619 | *** |

**Table S26:** Estimates for infection growth in unlock phase 1 using ITR.

|  | Estimate | 2.5_ci | 97.5_ci | SE | DF | T-stat | P-val | Sig |
| --- | --- | --- | --- | --- | --- | --- | --- | --- |
| Intercept | 0.16683821 | 0.12767608 | 0.20600033 | 0.01997613 | 4924 | 8.35187754 | 2.22E-16 | *** |
| days0 | -0.0037606 | -0.0048654 | -0.0026558 | 0.00056356 | 4924 | -6.6729745 | 2.78E-11 | *** |
| days1 | 0.00363807 | 0.00249789 | 0.00477826 | 0.00058159 | 4924 | 6.25536356 | 4.30E-10 | *** |
| phase_indi_4 | 0.00919676 | 0.00078048 | 0.01761303 | 0.00429304 | 4924 | 2.14224652 | 0.03222255 | * |
| log_sum_pop | -0.0037004 | -0.0057541 | -0.0016468 | 0.00104753 | 4924 | -3.532555 | 0.00041534 | *** |

**Table S27:** Estimates for infection growth in unlock phase 2 using ITR.

|  | Estimate | 2.5_ci | 97.5_ci | SE | DF | T-stat | P-val | Sig |
| --- | --- | --- | --- | --- | --- | --- | --- | --- |
| Intercept | 0.16845385 | 0.12932119 | 0.20758651 | 0.0199611 | 4924 | 8.43910667 | 0 | *** |
| days0 | -0.0039642 | -0.0050746 | -0.0028538 | 0.00056642 | 4924 | -6.9986791 | 2.93E-12 | *** |
| days1 | 0.00396428 | 0.00281031 | 0.00511824 | 0.00058862 | 4924 | 6.73481163 | 1.83E-11 | *** |
| phase_indi_5 | -0.0195162 | -0.0293876 | -0.0096447 | 0.00503532 | 4924 | -3.8758584 | 0.00010763 | *** |
| log_sum_pop | -0.0037004 | -0.0057519 | -0.001649 | 0.00104642 | 4924 | -3.5362919 | 0.00040952 | *** |

**Table S28:** Estimates for mobility in lockdown phase 1 using ITR.

|  | Estimate | 2.5_ci | 97.5_ci | SE | DF | T-stat | P-val | Sig |
| --- | --- | --- | --- | --- | --- | --- | --- | --- |
| (Intercept) | 76.5108973 | 52.8094603 | 100.212334 | 12.0927921 | 30.8116151 | 6.3269836 | 4.99E-07 | *** |
| days0 | -6.9840876 | -7.135934 | -6.8322412 | 0.07747407 | 4521.8231 | -90.147416 | 0 | *** |
| days1 | 7.17595408 | 7.02352626 | 7.32838191 | 0.07777073 | 4865.54259 | 92.2706298 | 0 | *** |
| phase_indi_0 | -8.0232295 | -8.7501714 | -7.2962877 | 0.3708955 | 4865.54029 | -21.632049 | 3.69E-99 | *** |
| log_sum_pop | -3.3323715 | -4.7355444 | -1.9291986 | 0.7159177 | 30.3755511 | -4.6546852 | 6.01E-05 | *** |

**(a) Fixed effects**
|  | Name | Var | Std |
| --- | --- | --- | --- |
| state | (Intercept) | 49.074794 | 7.00534039 |
| state | days0 | 0.00503324 | 0.07094535 |
| Residual |  | 51.981437 | 7.20981532 |

**Table S29:**
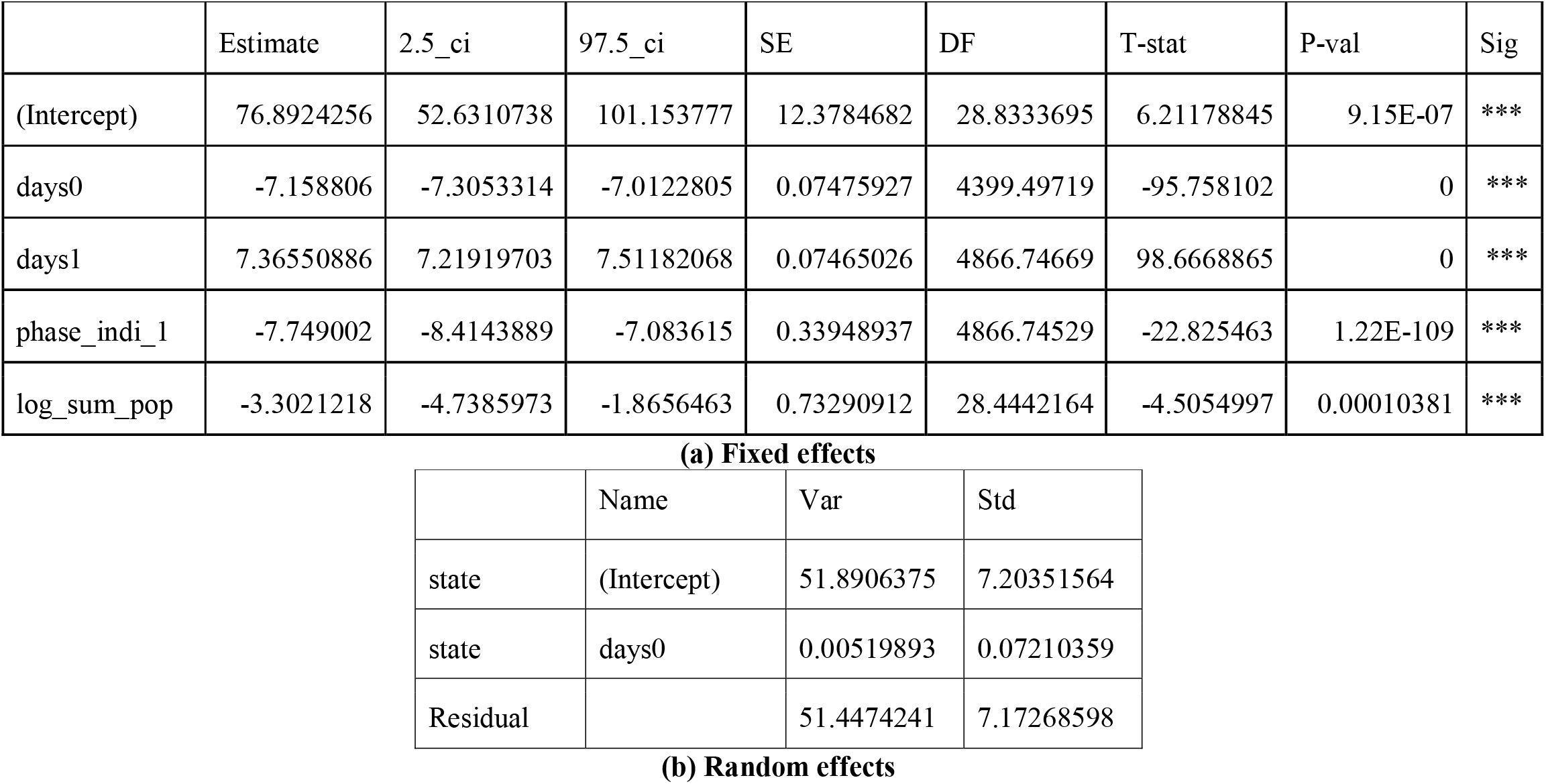
Estimates for mobility in lockdown phase 2 using ITR.

|  | Estimate | 2.5_ci | 97.5_ci | SE | DF | T-stat | P-val | Sig |
| --- | --- | --- | --- | --- | --- | --- | --- | --- |
| (Intercept) | 76.8924256 | 52.6310738 | 101.153777 | 12.3784682 | 28.8333695 | 6.21178845 | 9.15E-07 | *** |
| days0 | -7.158806 | -7.3053314 | -7.0122805 | 0.07475927 | 4399.49719 | -95.758102 | 0 | *** |
| days1 | 7.36550886 | 7.21919703 | 7.51182068 | 0.07465026 | 4866.74669 | 98.6668865 | 0 | *** |
| phase_indi_1 | -7.749002 | -8.4143889 | -7.083615 | 0.33948937 | 4866.74529 | -22.825463 | 1.22E-109 | *** |
| log_sum_pop | -3.3021218 | -4.7385973 | -1.8656463 | 0.73290912 | 28.4442164 | -4.5054997 | 0.00010381 | *** |

**(a) Fixed effects**
|  | Name | Var | Std |
| --- | --- | --- | --- |
| state | (Intercept) | 51.8906375 | 7.20351564 |
| state | days0 | 0.00519893 | 0.07210359 |
| Residual |  | 51.4474241 | 7.17268598 |

**Table S30:** Estimates for mobility in lockdown phase 3 using ITR.

|  | Estimate | 2.5_ci | 97.5_ci | SE | DF | T-stat | P-val | Sig |
| --- | --- | --- | --- | --- | --- | --- | --- | --- |
| (Intercept) | 75.1901979 | 52.1235067 | 98.256889 | 11.7689362 | 9.546751 | 6.38886952 | 9.78E-05 | *** |
| days0 | -7.5456379 | -7.6972562 | -7.3940195 | 0.07735774 | 4461.55083 | -97.542125 | 0 | *** |
| days1 | 7.7749637 | 7.62365185 | 7.92627555 | 0.07720134 | 4866.91944 | 100.710213 | 0 | *** |
| phase_indi_2 | 0.90070316 | 0.14246048 | 1.65894584 | 0.38686562 | 4866.91935 | 2.32820679 | 0.01994189 | * |
| log_sum_pop | -3.1077725 | -4.4830996 | -1.7324453 | 0.70171043 | 13.1546894 | -4.4288532 | 0.00066187 | *** |

**(a) Fixed effects**
|  | Name | Var | Std |
| --- | --- | --- | --- |
| state | (Intercept) | 132.563921 | 11.5136406 |
| state | days0 | 0.00520765 | 0.07216408 |
| state | log_sum_pop | 0.33289743 | 0.57697265 |
| Residual |  | 56.8871865 | 7.54235948 |

**Table S31:** Estimates for mobility in lockdown phase 4 using ITR.

|  | Estimate | 2.5_ci | 97.5_ci | SE | DF | T-stat | P-val | Sig |
| --- | --- | --- | --- | --- | --- | --- | --- | --- |
| (Intercept) | 78.4815054 | 55.0502713 | 101.912739 | 11.954931 | 11.2303471 | 6.56478115 | 3.67E-05 | *** |
| days0 | -7.6227958 | -7.7709515 | -7.4746401 | 0.07559103 | 4410.65921 | -100.84259 | 0 | *** |
| days1 | 7.85395638 | 7.70627022 | 8.00164253 | 0.07535146 | 4865.19645 | 104.230973 | 0 | *** |
| phase_indi_3 | 5.28743784 | 4.55429977 | 6.0205759 | 0.37405691 | 4863.88295 | 14.1353835 | 1.73E-44 | *** |
| log_sum_pop | -3.2585928 | -4.6535036 | -1.863682 | 0.71170228 | 12.3547924 | -4.5785898 | 0.00058867 | *** |

**(a) Fixed effects**
|  | Name | Var | Std |
| --- | --- | --- | --- |
| state | (Intercept) | 43.805225 | 6.61855158 |
| state | days0 | 0.00519954 | 0.07210786 |
| state | log_sum_pop | 0.06266365 | 0.25032709 |
| Residual |  | 54.7117679 | 7.39674036 |

**Table S32:**
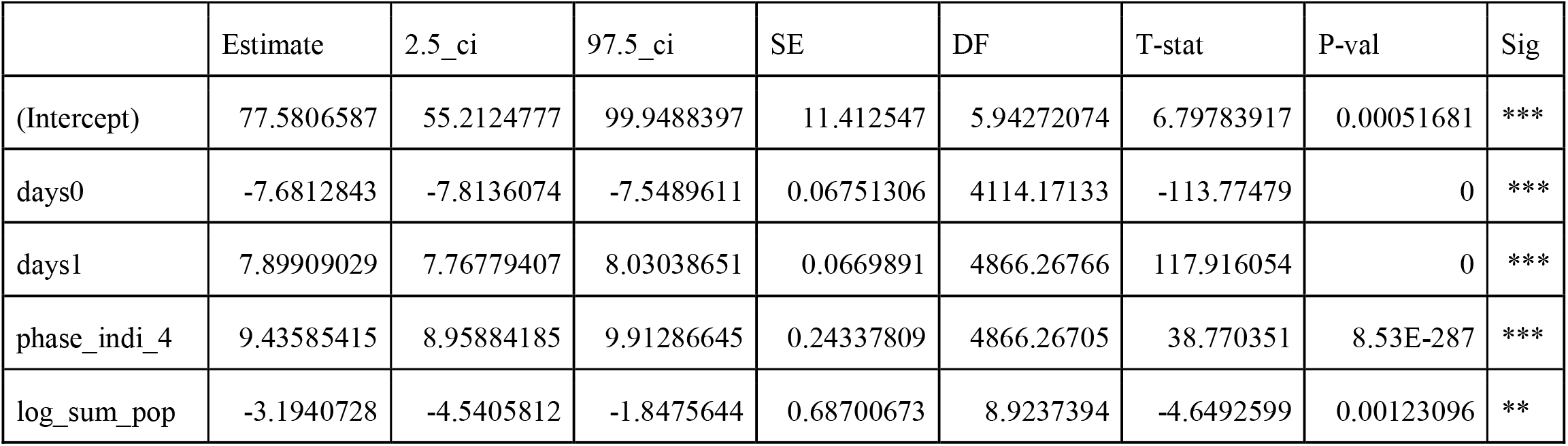
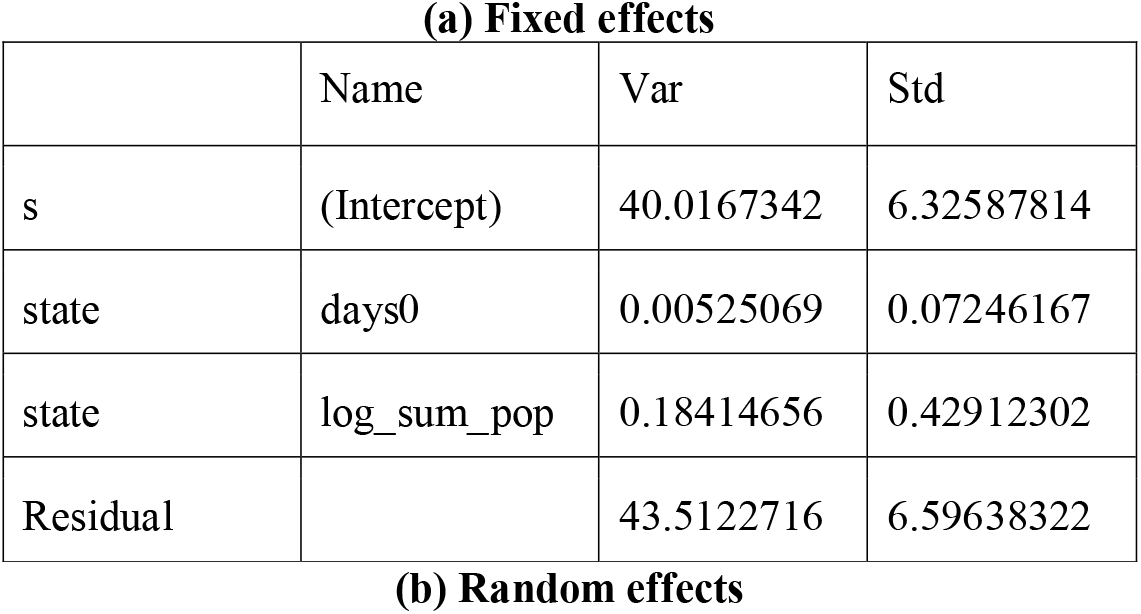
Estimates for mobility in unlock phase 1 using ITR.

**Table S33:** Estimates for mobility in unlock phase 2 using ITR.

**(b) Random effects****Table S33: Estimates for mobility in unlock phase 2 using ITR**
|  | Estimate | 2.5_ci | 97.5_ci | SE | DF | T-stat | P-val | Sig |
| --- | --- | --- | --- | --- | --- | --- | --- | --- |
| (Intercept) | 80.4308751 | 58.1703532 | 102.691397 | 11.3576179 | 1.39613775 | 7.08166767 | 0.04621207 | * |
| days0 | -7.5549238 | -7.7051071 | -7.4047405 | 0.07662555 | 4383.86181 | -98.595363 | 0 | *** |
| days1 | 7.79176613 | 7.64186568 | 7.94166659 | 0.07648123 | 4862.80877 | 101.878149 | 0 | *** |
| phase_indi_5 | -1.9473124 | -2.5541829 | -1.340442 | 0.30963347 | 4862.95788 | -6.289089 | 3.47E-10 | *** |
| log_sum_pop | -3.3691215 | -4.7314158 | -2.0068272 | 0.69506088 | 3.60705381 | -4.8472322 | 0.01080497 | * |

**(a) Fixed effects**
|  | Name | Var | Std |
| --- | --- | --- | --- |
| state | (Intercept) | 18.6881432 | 4.32297851 |
| state | days0 | 0.00530359 | 0.07282573 |
| state | log_sum_pop | 0.49104982 | 0.70074947 |
| Residual |  | 56.4805422 | 7.51535377 |

**Table S34:** Estimates for Rt in lockdown phase 1 using ITR.

**(b) Random effects****Table S34: Estimates for Rt in lockdown phase 1 using ITR**
|  | Estimate | 2.5_ci | 97.5_ci | SE | DF | T-stat | P-val | Sig |
| --- | --- | --- | --- | --- | --- | --- | --- | --- |
| (Intercept) | 1.71001543 | -0.056759 | 3.47678989 | 0.9014321 | 33.8834924 | 1.89699859 | 0.06637847 | . |
| days0 | -0.0550437 | -0.0884921 | -0.0215953 | 0.01706584 | 2095.23665 | -3.2253735 | 0.00127746 | ** |
| days1 | 0.0458001 | 0.01247108 | 0.07912913 | 0.01700492 | 4865.78651 | 2.6933446 | 0.00709817 | ** |
| phase_indi_0 | -0.4464996 | -0.7538205 | -0.1391787 | 0.15679926 | 4865.78664 | -2.8475875 | 0.00442366 | ** |
| log_sum_pop | 0.08778449 | -0.0316673 | 0.20723628 | 0.06094591 | 45.8942969 | 1.44036722 | 0.15655107 |  |

**(a) Fixed effects**
|  | Name | Var | Std |
| --- | --- | --- | --- |
| state | days0 | 0.00080495 | 0.02837156 |
| state | log_sum_pop | 0.03044006 | 0.1744708 |
| Residual |  | 9.36572491 | 3.06034719 |

**Table S35:** Estimates for Rt in lockdown phase 2 using ITR.

**(b) Random effects****Table S35: Estimates for Rt in lockdown phase 2 using ITR**
|  | Estimate | 2.5_ci | 97.5_ci | SE | DF | T-stat | P-val | Sig |
| --- | --- | --- | --- | --- | --- | --- | --- | --- |
| (Intercept) | 1.91982802 | 0.1630553 | 3.67660074 | 0.89632908 | 34.2696102 | 2.14187853 | 0.03939403 | * |
| days0 | -0.0894392 | -0.1214071 | -0.0574712 | 0.01631047 | 2066.94061 | -5.4835427 | 4.68E-08 | *** |
| days1 | 0.08396506 | 0.05244071 | 0.11548941 | 0.01608415 | 4864.84595 | 5.22036093 | 1.86E-07 | *** |
| phase_indi_1 | 0.60148115 | 0.319181 | 0.8837813 | 0.14403334 | 4864.84574 | 4.17598568 | 3.02E-05 | *** |
| log_sum_pop | 0.09057457 | -0.0275503 | 0.20869948 | 0.06026892 | 46.4628795 | 1.50284059 | 0.13964706 |  |

**(a) Fixed effects**
|  | Name | Var | Std |
| --- | --- | --- | --- |
| state | days0 | 0.00076814 | 0.02771538 |
| state | log_sum_pop | 0.02882982 | 0.16979346 |
| Residual |  | 9.35106061 | 3.05795039 |

**Table S36:**
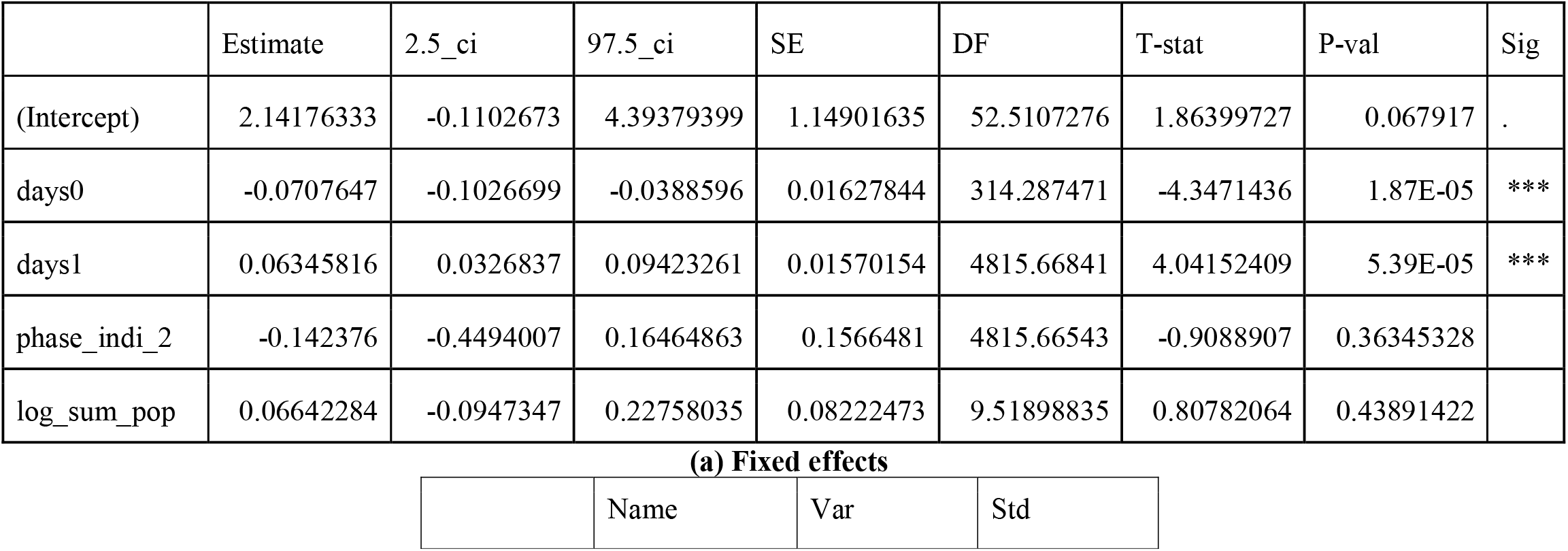
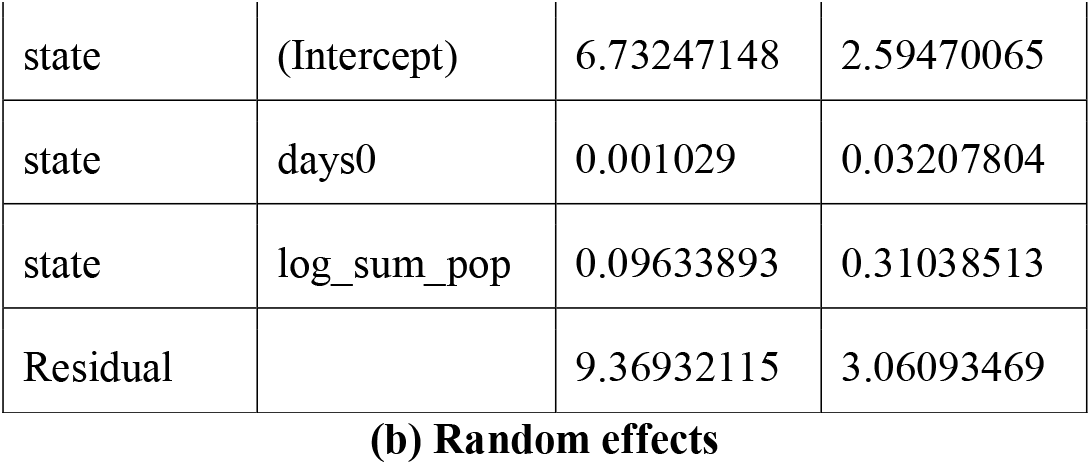
Estimates for Rt in lockdown phase 3 using ITR.

**(b) Random effects****Table S36: Estimates for Rt in lockdown phase 3 using ITR**
|  | Estimate | 2.5_ci | 97.5_ci | SE | DF | T-stat | P-val | Sig |
| --- | --- | --- | --- | --- | --- | --- | --- | --- |
| (Intercept) | 2.14176333 | -0.1102673 | 4.39379399 | 1.14901635 | 52.5107276 | 1.86399727 | 0.067917 | . |
| days0 | -0.0707647 | -0.1026699 | -0.0388596 | 0.01627844 | 314.287471 | -4.3471436 | 1.87E-05 | *** |
| days1 | 0.06345816 | 0.0326837 | 0.09423261 | 0.01570154 | 4815.66841 | 4.04152409 | 5.39E-05 | *** |
| phase_indi_2 | -0.142376 | -0.4494007 | 0.16464863 | 0.1566481 | 4815.66543 | -0.9088907 | 0.36345328 |  |
| log_sum_pop | 0.06642284 | -0.0947347 | 0.22758035 | 0.08222473 | 9.51898835 | 0.80782064 | 0.43891422 |  |

**(a) Fixed effects**
|  | Name | Var | Std |
| --- | --- | --- | --- |
| state | (Intercept) | 6.73247148 | 2.59470065 |
| state | days0 | 0.001029 | 0.03207804 |
| state | log_sum_pop | 0.09633893 | 0.31038513 |
| Residual |  | 9.36932115 | 3.06093469 |

**Table S37:** Estimates for Rt in lockdown phase 4 using ITR.

|  | Estimate | 2.5_ci | 97.5_ci | SE | DF | T-stat | P-val | Sig |
| --- | --- | --- | --- | --- | --- | --- | --- | --- |
| (Intercept) | 3.60078298 | 1.9328885 | 5.26867746 | 0.85098221 | 50.7164293 | 4.2313258 | 9.74E-05 | *** |
| days0 | -0.0677994 | -0.0980909 | -0.037508 | 0.0154551 | 4816.93259 | -4.3868616 | 1.17E-05 | *** |
| days1 | 0.06045231 | 0.0297078 | 0.09119681 | 0.01568626 | 4823.84193 | 3.85383794 | 0.00011779 | *** |
| phase_indi_3 | -0.486859 | -0.7921052 | -0.1816128 | 0.15574072 | 4823.84164 | -3.126087 | 0.00178202 | ** |
| log_sum_pop | -0.0131414 | -0.1004147 | 0.07413185 | 0.04452799 | 30.3148614 | -0.2951269 | 0.76990881 |  |

**(a) Fixed effects**
|  | Name | Var | Std |
| --- | --- | --- | --- |
| state | (Intercept) | 3.06288388 | 1.75010968 |
| state | days0 | 0.000245 | 0.01565258 |
| Residual |  | 9.48749588 | 3.0801779 |

**Table S38:** Estimates for Rt in unlock phase 1 using ITR.

|  | Estimate | 2.5_ci | 97.5_ci | SE | DF | T-stat | P-val | Sig |
| --- | --- | --- | --- | --- | --- | --- | --- | --- |
| (Intercept) | 2.02139615 | 0.18348268 | 3.85930962 | 0.93772819 | 29.3448459 | 2.15563122 | 0.03944449 | * |
| days0 | -0.0739399 | -0.1063158 | -0.041564 | 0.01651862 | 325.727527 | -4.4761541 | 1.05E-05 | *** |
| days1 | 0.06655263 | 0.03618341 | 0.09692186 | 0.01549479 | 4869.744 | 4.29516265 | 1.78E-05 | *** |
| phase_indi_4 | 0.14011907 | -0.0840522 | 0.36429033 | 0.11437519 | 4869.74452 | 1.22508277 | 0.22060329 |  |
| log_sum_pop | 0.07740518 | -0.0581349 | 0.21294522 | 0.06915435 | 31.627796 | 1.11931027 | 0.27143666 |  |

**(a) Fixed effects**
|  | Name | Var | Std |
| --- | --- | --- | --- |
| state | days0 | 0.00142266 | 0.03771821 |
| state | log_sum_pop | 0.05493586 | 0.234384 |
| Residual |  | 9.35028423 | 3.05782345 |

**Table S39:** Estimates for Rt in unlock phase 2 using ITR.

|  | Estimate | 2.5_ci | 97.5_ci | SE | DF | T-stat | P-val | Sig |
| --- | --- | --- | --- | --- | --- | --- | --- | --- |
| (Intercept) | 5.60079684 | 2.15797143 | 9.04362224 | 1.75657585 | 569.931131 | 3.18847423 | 0.00150872 | ** |
| days0 | -0.0739711 | -0.1052512 | -0.042691 | 0.01595954 | 1633.12416 | -4.6349135 | 3.85E-06 | *** |
| days1 | 0.06723231 | 0.03645457 | 0.09801005 | 0.01570322 | 4887.41387 | 4.28143587 | 1.89E-05 | *** |
| phase_indi_5 | -0.0761629 | -0.3394472 | 0.18712131 | 0.13433117 | 4887.41375 | -0.566979 | 0.57075451 |  |
| log_sum_pop | -0.1271956 | -0.3184243 | 0.06403312 | 0.09756747 | 57.2145961 | -1.3036682 | 0.19756804 |  |

**(a) Fixed effects**
|  | Name | Var | Std |
| --- | --- | --- | --- |
| state | (Intercept) | 44.696988 | 6.6855806 |
| state | days0 | 0.00079065 | 0.02811857 |
| state | log_sum_pop | 0.13553002 | 0.36814402 |
| Residual |  | 9.35563387 | 3.05869807 |

## Notes

### Competing Interest Statement

The authors have declared no competing interest.

